# FRANK: a pan-cancer RNA-seq classifier for childhood tumors with data-efficient learning

**DOI:** 10.64898/2026.09.23.26355605

**Authors:** Karolis Sablauskas, Tatjana Kiselova, Livija Bardina, Inga Nartisa, Andrius Zucenka, Agnese Viluma, Linda Gailite, Audrone Jakaitiene, Egija Berga-Svitina, Dmitrijs Rots

## Abstract

Accurate characterization of tumor subtype is of paramount importance in guiding treatment decisions in pediatric cancers. Here, we present FRANK (Fully-connected RNA-based Augmentation Neural Klassifier), a machine learning approach for classifying pan-cancer pediatric tumors. We curated a set of 11,467 transcriptomes to build a high-quality training dataset representing 181 distinct childhood tumor and normal tissue types. We used extensive data augmentation techniques with a hierarchical penalty term to train a neural network capable of detecting tumor types with as few as three training examples. To evaluate the generalization of the trained model, we perform validation on nine datasets spanning 16,398 samples, achieving an overall accuracy of 98.3%. Furthermore, we demonstrated that FRANK outperformed other published pediatric pan-cancer RNA-seq classifiers on an external dataset. To evaluate the robustness, we showed that FRANK remained accurate when applied to long-read RNA-sequencing data, as well as to noisy and low-quality data.

## Main

In developed countries, cancer remains the leading cause of death by disease in children. Over the last 40 years, the incidence of pediatric tumors has increased based on registry data from several EU countries^1^. With an increasing number of available therapeutic options and risk-adjusted protocols, accurate diagnostics remain a fundamental step in treating pediatric malignancies^2^. The number of subclasses of childhood tumors is increasing over time, in part due to advances in sequencing technologies, and increasingly relies on disease entity definition according to hallmark genomic aberrations requiring multiple tests, including multi-omic analysis and advanced bioinformatics resources^2,3^. For example, Heidelberg CNS Tumor Methylation Classifier expanded the number of recognized disease entities from 91 to 184 subclasses^4^. While some childhood tumors such as Wilms tumor or B-cell acute lymphoblastic leukemia (B-ALL) are more common in the pediatric population – all pediatric tumor types are considered rare disorders (with prevalence <1:2,000)^5^. Additionally, rare tumor subtypes collectively comprise a sizable proportion of all pediatric cancers, e.g., tumors with an annual incidence <2/1,000,000 corresponded to 11% of all cancers in patients aged 0–14 years^6^.

Whole transcriptome sequencing (WTS)/RNA-seq has recently emerged as a method for capturing a diverse set of genomic changes^7,8^, including fusions, splice variants, and even chromosomal number aberrations. However, detecting these changes requires specialized bioinformatics tools, usually suited to a single type of analysis^8^. Additionally, running this kind of analysis can be computationally expensive. Since different tumor types have distinct gene expression profiles, this can be used to classify tumors.

However, the training of classification models requires a large training set, which is difficult to obtain for rare disorders like pediatric cancers. Nonetheless, pan-cancer classifiers using transcriptomes have been described for pediatric tumors. For instance, OTTER^9^ is a one-dimensional convolutional neural network-based approach for pan-cancer classification. Recently, also M&M^10^, a majority-minority classifier based on the random forest (RF) and *k*-nearest neighbors classifier that can identify 96 pediatric tumor subtypes, has been described. Both classifiers had limited testing on external data, and although the number of tumor types detected was high in both cases, some newly emerging tumor categories, such as CNS tumor and T-ALL subclasses, were absent.

In this study, we describe the development and validation of a transcriptome-based pan-cancer classifier designated FRANK (Fully-connected RNA-based Augmentation Neural Klassifier), specialized for pediatric and young adult-onset tumors, including rare subclasses. We show its suitability for clinical application based on comprehensive testing in independent cohorts and different clinical scenarios.

## Results

To address the limited external testing, generalizability analysis and representation of emerging pediatric tumor classes, we developed a fine-level pediatric pan-cancer RNA classifier that was extensively validated on external and out-of-distribution data.

First, we curated data from pediatric pan-cancer cohorts (St. Jude Children’s Research Hospital, USA^11^ and Princess Maxima Center for Pediatric Oncology, The Netherlands^10^) with detailed molecular descriptions/classes. To support the detection of rare entities, we supplemented the training dataset with T-ALL cases from the AALL0434 study^12^. Finally, to differentiate normal from malignant tissues, we supplemented the training cohort with healthy tissue samples from the Genotype-Tissue Expression project (GTEx)^13^ (Fig. 1). Extensive quality control (QC) was performed to exclude low-quality samples from the training cohort. In total, this yielded 5,714 tumor and 5,753 normal tissue samples (total *n=*11,467) from 5,615 individuals (Table 1).

**Fig. 1:**
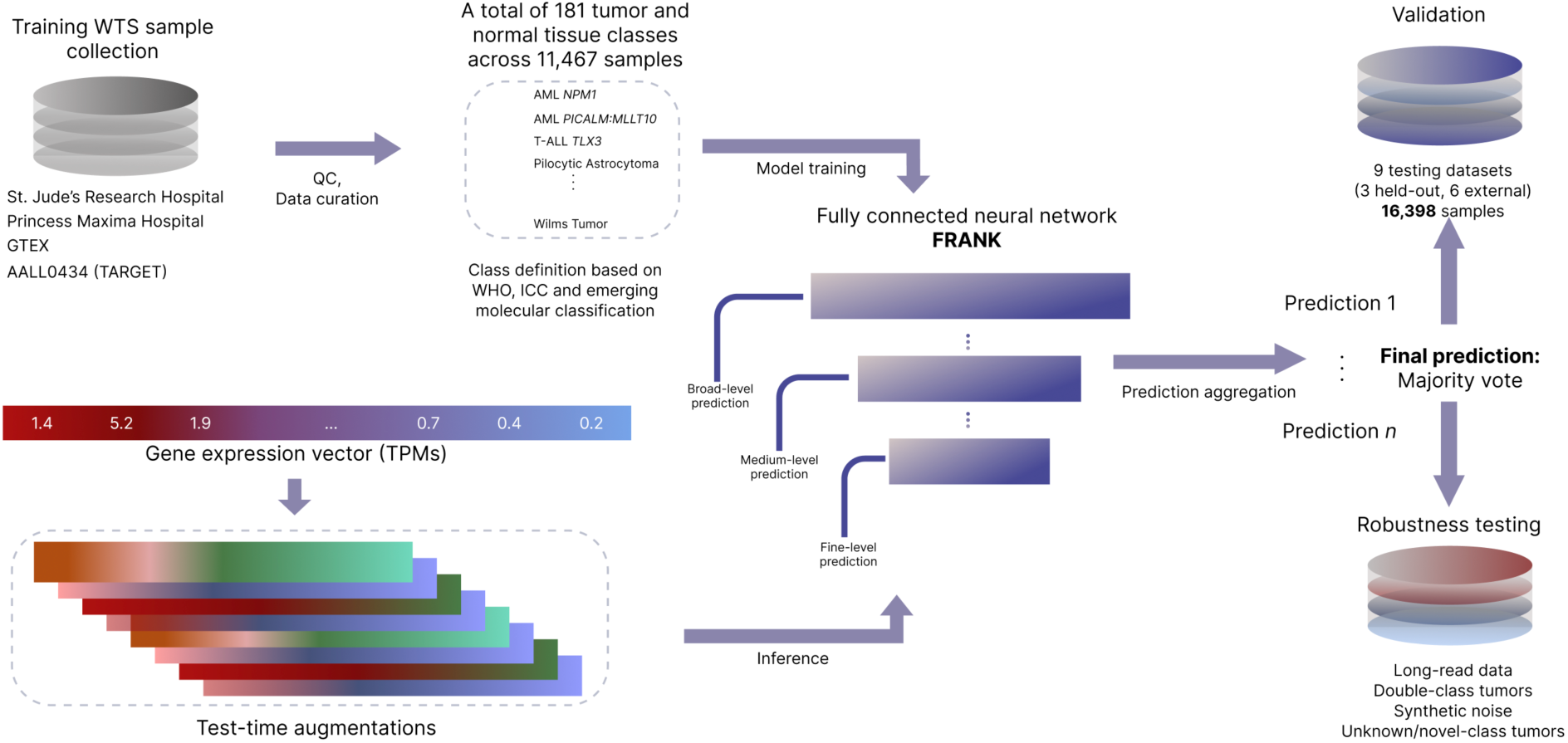
Overview of training and testing of FRANK. Starting from top left: The training samples were sourced from four datasets. Following QC and data curation, a total of 181 tumor and normal tissue classes were defined. The model was trained on a total of 11,467 samples and then tested on nine datasets, six of which were fully independent from training datasets. FRANK is a three-layer fully connected neural network where each layer outputs a prediction for a progressively finer tumor hierarchy level. The model’s robustness and generalization were evaluated on a set of out-of-distribution samples, including long-read data, tumors with more than one hallmark molecular aberration, samples with artificial noise and low-quality samples. For inference, samples were processed as a normalized gene expression vector with each sample being processed 20 times using test-time augmentations. The final prediction corresponded to a majority vote across these predictions. TPMs – transcripts per million.

**Table 1:** Training cohort summary.

| Cohort | N Individual<br>s | N Samples | N Classes<br>(fine level) | Tissue<br>preservative | Enrichment<br>method | GENCODE<br>version |
| --- | --- | --- | --- | --- | --- | --- |
| St. Jude | 3,911 | 4,319<br>(Tumor<br>*n*=4319)<br>(Normal<br>*n*=0) | 144 (Pan-<br>tumor) | FFPE (*n*=805)<br>Fresh/Stabilized<br>(*n*=2,973)<br>Not Available<br>(*n*=541) | Not Available (*n*=5)<br>PolyA (*n*=889) Total<br>RNA (*n*=3,425) | v31 |
| Princess Maxima | 1,136 | 1,149<br>(Tumor<br>*n*=1110)<br>(Normal<br>*n*=39) | 80 (Pan-<br>tumor) | Fresh/Stabilized<br>(*n*=1149) | Total RNA<br>(*n*=1,149) | v29 |
| GTEX v10<br>release | 283 | 5,714<br>(Tumor<br>*n*=0)<br>(Normal<br>*n*=5,714) | 31 (All<br>normal tissue) | Fresh/Stabilized<br>(*n*=5714) | Total RNA<br>(*n*=5,714) | v39 |
| AALL0434 | 285 | 285 (Tumor<br>*n*=285)<br>(Normal<br>*n*=0) | 7 (T-ALL) | Fresh/Stabilized<br>(*n*=285) | Total RNA (*n*=285) | N.A. |
| Total | 5,615 | 11,467<br>(Tumor<br>*n*=5,714)<br>(Normal<br>*n*=5,753) | 181 |  |  |  |

*Summary of training samples used in training. FFPE - formalin-fixed paraffin-embedded. T-ALL - T-cell* *acute lymphoblastic leukemia.*

All samples were re-evaluated based on the provided molecular findings/classes and assigned to three levels of tumor hierarchy which after data curation, resulted in three broad-level categories (hematologic, solid and brain), 48 medium-level classes and 181 fine-level classes. We aimed to assign the samples to a detailed molecular subclass, based on the confirmed and emerging World Health Organization v5 (WHO) and/or International Consensus Classification (ICC) entities or to other molecularly defined categories. To be able to detect rare tumor types, we set the lowest number of training examples per fine-level class to three.

Next, we selected a set of genes to be used as the model input by first training an RF classifier using a three-fold split (corresponding to the lowest number of training samples per class) and then using the resulting feature importance to select the top 7,000 genes. TPM values were log_2_-normalized and genes with variance below 1.0 were filtered out, yielding a total of 5,260 genes to be used for the model input. Standard scaling was applied prior to model input.

We then trained a neural network model that could leverage the hierarchical classification of tumor categories given that high-level classes are easier to predict than fine-level ones. To achieve this, we introduced a differentiable hierarchical penalty term in training and structured the model to output broad-, medium– and fine-level class predictions in successive model layers. To maximize the use of data from rare classes, we generated synthetic samples for classes with n≤5 and trained on all training samples. The use of train-time augmentations, in particular CutMix^14^, allowed us to counteract overfitting and train the model for more than 100 epochs without the use of a hold-out dataset. For inference, 20 test-time augmentations were used for each sample. We designated the final trained model FRANK (**F**ully-connected **R**NA-based **A**ugmentation **K**lassifier) to reflect the extensive use of data augmentation techniques. The model outputs the top three predictions for fine-level classes and a prediction for medium and broad-level classes with prediction confidence scores to support the interpretation of the results. For comparison to other widely used machine learning (ML) approaches, we trained logistic regression (LR), RF, support vector machine (SVM) and extreme gradient boosting (XGB) classifiers to predict fine-level classes using the same training dataset.

### Validation on tumor samples

To evaluate FRANK’s performance, we focused on testing the model on external tumor datasets. For this, we sourced a pan-cancer tumor dataset from Riga Children’s Clinical University Hospital (RCCUH), which had not been used in any previously described models. The dataset contained 178 tumor samples with fine-level molecular classes and FRANK achieved 92.7% accuracy and an F1 score of 0.87 (Table 2). Next, we tested FRANK on 430 held-out malignant samples from Princess Maxima Center for Pediatric Oncology^10^ and achieved a comparable performance with an accuracy of 95.1% and an F1 score of 0.90 (Table 2). Further, we expanded the testing to the Therapeutically Applicable Research to Generate Effective Treatments (TARGET) cohorts which included *n*=489 acute myeloid leukemia (AML) samples (TARGET AML)^15^ and a set of *n*=468 brain and solid tumors (TARGET Other)^16–19^, as well as *n*=707 held-out T-ALL samples (AALL0434^12^). Again, FRANK delivered a high performance across all three datasets, with accuracies of 91.4%, 93.2% and 94.8% and F1 scores of 0.80, 0.90 and 0.85 for TARGET AML, TARGET Other and AALL0434, respectively (Table 2). For additional testing on T-ALL samples, an external cohort from Saint-Louis Hospital^20^, France (*n=*72) was used. The performance on this cohort was comparable to that on the held-out AALL0434 samples, with an accuracy of 90.3% and an F1 score of 0.87 (Table 2). Finally, to evaluate the model’s performance on a cohort with disjoint patient characteristics but similar disease entities, we ran FRANK on the Beat AML cohort^21^, which included adult subjects with AML or myelodysplastic syndrome. Of 735 Beat AML cases, 329 (44.8%) had a defining molecular change, of which 280 were classified correctly, resulting in an accuracy of 85.1% and an F1 score of 0.72 (Table 2). Regarding medium-level predictions on the Beat AML cohort, 683/735 (92.9%) of cases were predicted as AML.

**Table 2:**
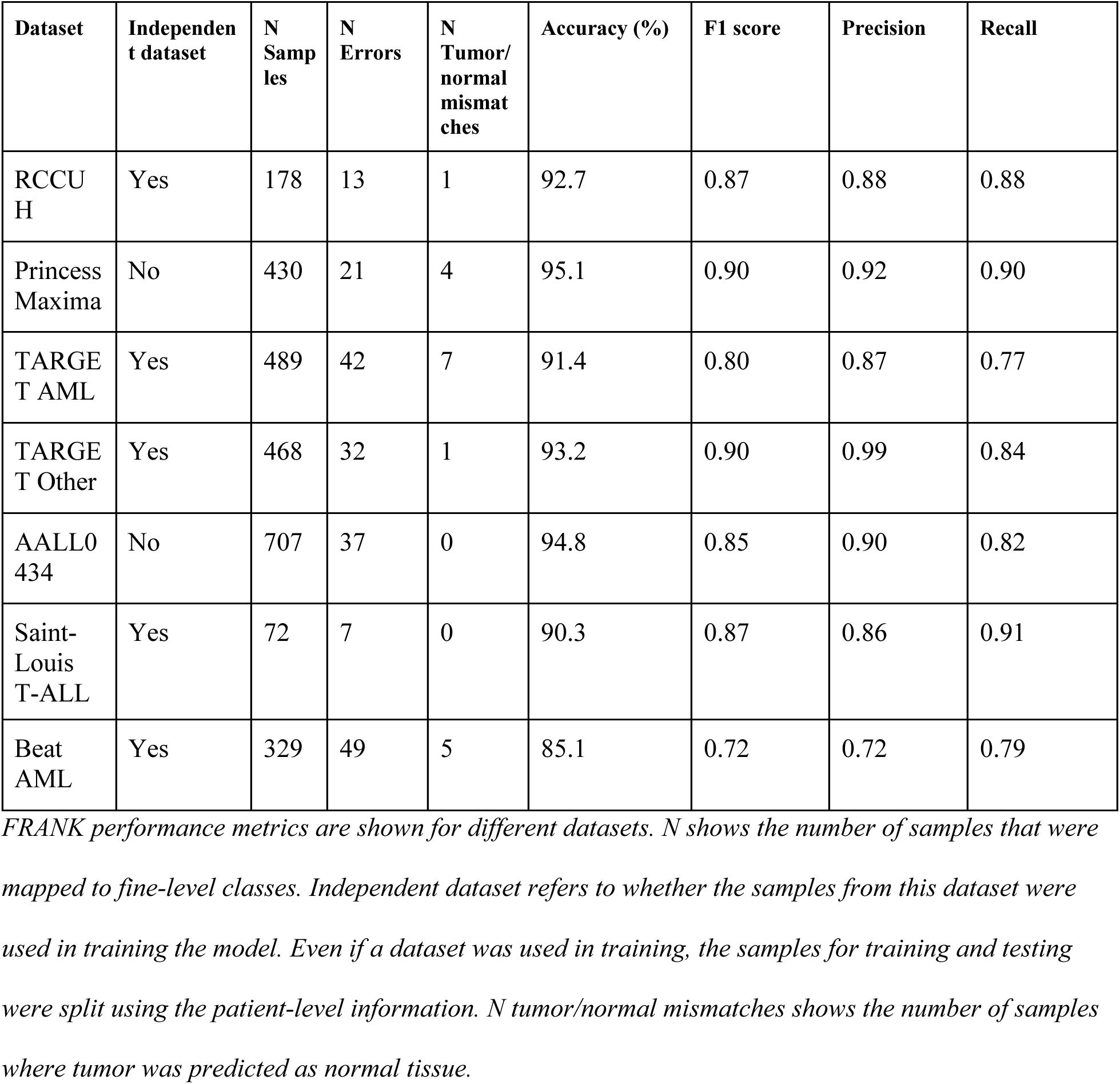
Performance of FRANK on tumor samples across validation datasets.

### Validation on normal tissue

To evaluate the performance also on samples without a known malignancy, we sourced a dataset from Riga Stradins University (RSU) that included whole blood samples of individuals with inborn errors of immunity and their parents (*n=*123)^22^. FRANK correctly classified 117 (95.1%) of these samples (F1 score of 0.98; Table 3), all incorrect predictions were “Myelodysplastic Syndromes” with low confidence scores (range 0.30–0.48). Additionally, 15/16 (93.8%) of normal tissue samples from the Princess Maxima hold-out testing dataset were classified correctly (F1 score of 0.97; Table 3). The one incorrect prediction was misclassification of a normal lymph node sample as Hodgkin lymphoma. All 11 normal tissue samples from RCCUH were correctly identified as normal tissue (accuracy of 100%; F1 score of 1.0; Table 3). Finally, the majority of held-out GTEx samples were also classified correctly as normal tissue (13,511/13,575), with an overall accuracy of 99.5% and an F1 score of 0.98 (Table 3). Thus, similar to the tumor datasets above, FRANK also demonstrated consistently high accuracy across normal tissue cohorts.

**Table 3:** Performance of FRANK on normal tissue samples across validation datasets.

| Dataset | Independent dataset | N Samples | N Errors | N Normal/tumor mismatches | Accuracy (%) | F1 score | Precision | Recall |
| --- | --- | --- | --- | --- | --- | --- | --- | --- |
| RSU | Yes | 123 | 6 | 6 | 95.1 | 0.98 | 1.00 | 0.95 |
| Princess Maxima | No | 16 | 1 | 1 | 93.8 | 0.97 | 1.00 | 0.95 |
| RCCUH | Yes | 11 | 0 | 0 | 100.0 | 1.00 | 1.00 | 1.00 |
| GTEx v10 release | No | 13575 | 64 | 1 | 99.5 | 0.98 | 0.98 | 0.98 |

*FRANK performance metrics are shown for different datasets. N shows the number of samples that were* *mapped to fine-level classes. Independent dataset refers to whether the samples from this dataset were* *used in training the model. Even if a dataset was used in training, the samples for training and testing* *were split using the patient-level information. N normal/tumor mismatches shows the number of samples* *where normal tissue was predicted as tumor.*

### Benchmarking against other classifiers

To compare FRANK to other classifiers, we benchmarked it against two published pediatric pan-cancer classifiers, namely, M&M^10^ and OTTER^9^. M&M supports only 96 fine-level pediatric tissue type classes, OTTER supports 234 fine-level classes (pediatric and adult), while FRANK supports 181 entity classifications. The primary benchmarking dataset was RCCUH because it had not been used for the training of other classifiers and represented a broad spectrum of comprehensively tested pediatric tumors. For comparison, we first filtered normal and malignant samples that could be mapped to available fine-level classes in the other classifiers which resulted in 172/189 (91.0%) and 157/189 (83.1%) samples mapping to M&M and OTTER, respectively. All 157 samples mapped to OTTER were also possible to assign to M&M classes. FRANK outperformed both classifiers, achieving an accuracy of 93.6% vs 84.3% with an F1 score of 0.88 vs 0.75 when compared to M&M (Fig. 2a) and an accuracy of 94.3% vs 67.5% with an F1 score of 0.89 vs 0.76 when compared to OTTER (Fig. 2b). In fact, FRANK’s top-1 prediction outperformed any of the top-3 predictions provided by M&M (Fig. 2a). In a three-model comparison, on 157 samples mapping to both OTTER and M&M, FRANK outperformed both classifiers, while OTTER had a higher F1 score, precision and recall compared to M&M (Fig. 2b). Since M&M used a different GENCODE version (v29) for its training samples, we additionally remapped the RCCUH samples to GENCODE v29 (RCCUH-29). This had very little effect on the results, with FRANK still achieving a higher performance (Extended Data Fig. 1h).

**Fig. 2:**
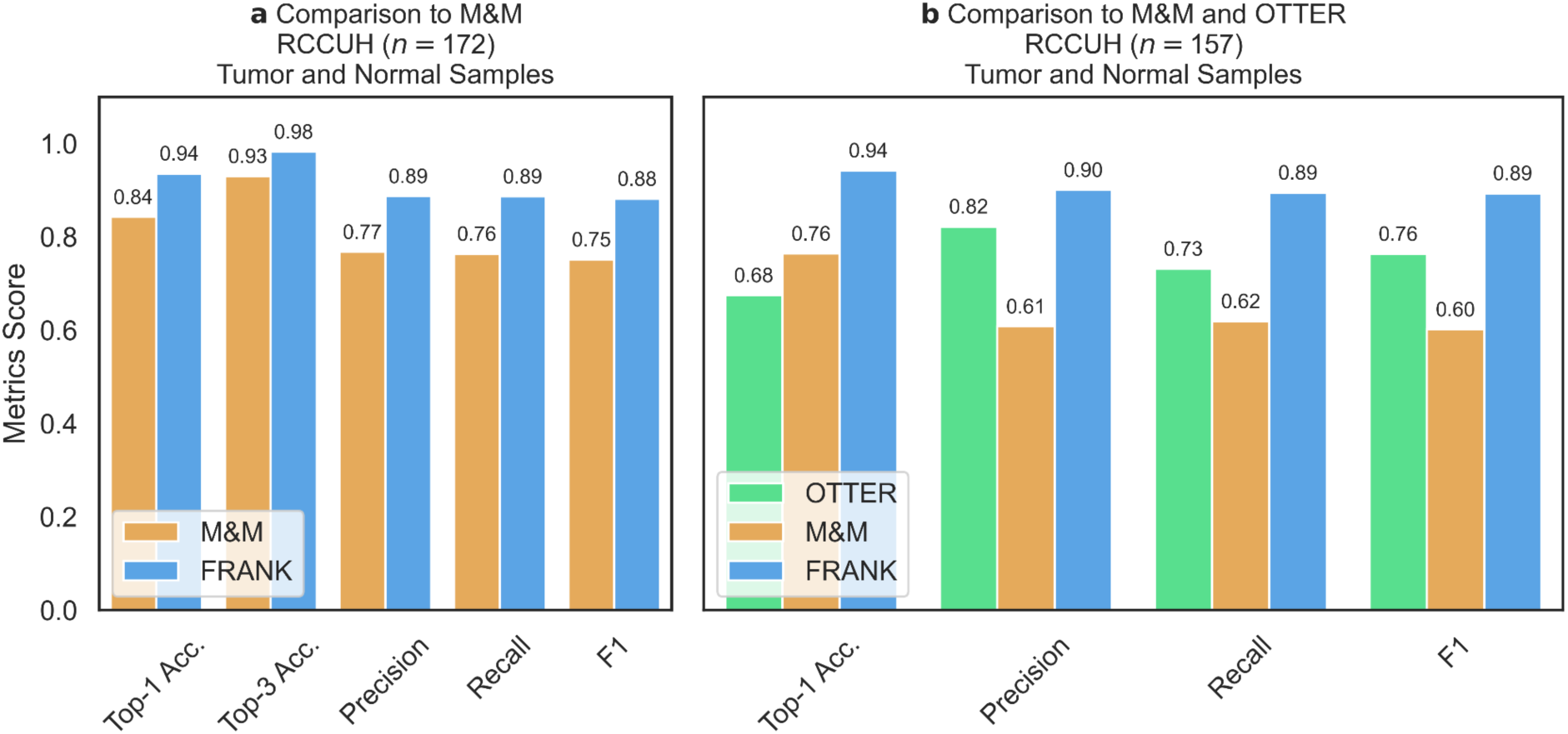
Benchmarking against other classifiers. **a**, Benchmarking of FRANK on RCCUH samples mapping to M&M classes. Not all samples mapping to M&M could additionally be mapped to OTTER, thus comparison of two tools is shown. **b**, Benchmarking of FRANK on RCCUH samples mapping to OTTER classes. All samples that are mapped to OTTER could additionally be mapped to M&M, thus comparison of three tools is shown. All metrics are shown in the range 0–1. Top-3 accuracy could not be extracted from OTTER and only top-1 accuracy is shown for comparison.

To perform a more comprehensive benchmarking, we extended the testing datasets. Since OTTER model weights are not publicly available, it was not possible to benchmark OTTER against other datasets. The M&M model is open-source and could be run on other datasets, with the exception of GTEx, because M&M supports only a limited number of normal tissues. Notably, FRANK outperformed M&M on the Princess Maxima testing cohort with accuracy of 95.1% vs. 89.0% and an F1 score of 0.89 vs 0.82 (Extended Data Fig. 1a). In all other cohorts, FRANK maintained and even increased this level of performance compared to M&M on cases that could be mapped to M&M-supported fine-level classes, whilst supporting almost double the number of classes (Extended Data Fig. 1b-h). Additionally, FRANK outperformed other ML models we have trained on the most tested datasets (Extended Data Fig. 2a-h)

### Aggregated analysis

We next aggregated the testing cohorts (RCCUH, AALL0434, GTEx, Princess Maxima, RSU, TARGET AML, TARGET Other, and Beat AML) into defined fine-level classes, yielding *n*=16,398 scorable samples to be used for overall performance assessment. The overall accuracy on the scorable dataset was 98.3% and the F1 score was 0.86 (Supplementary table 11), with an AUC of 0.96 (95% CI 0.94–0.97; Fig. 3a). We used the scorable test samples to perform sensitivity/specificity analysis based on confidence threshold (Fig. 3b). The optimal threshold was chosen as the point of intersection of sensitivity and specificity, corresponding to ∼0.95. Filtering by this threshold yielded 14,878 (90.7%) scorable samples and showed performance improvements across all metrics, with accuracy reaching 99.9% (Supplementary table 11), an F1 score of 0.99 and only 22 misclassified samples. The accuracy of high confidence predictions was similar between tumor samples and the normal tissues: 99.4% and 99.9% with an F1 score of 0.98 and 0.99, respectively (Supplementary table 11).

**Fig. 3:**
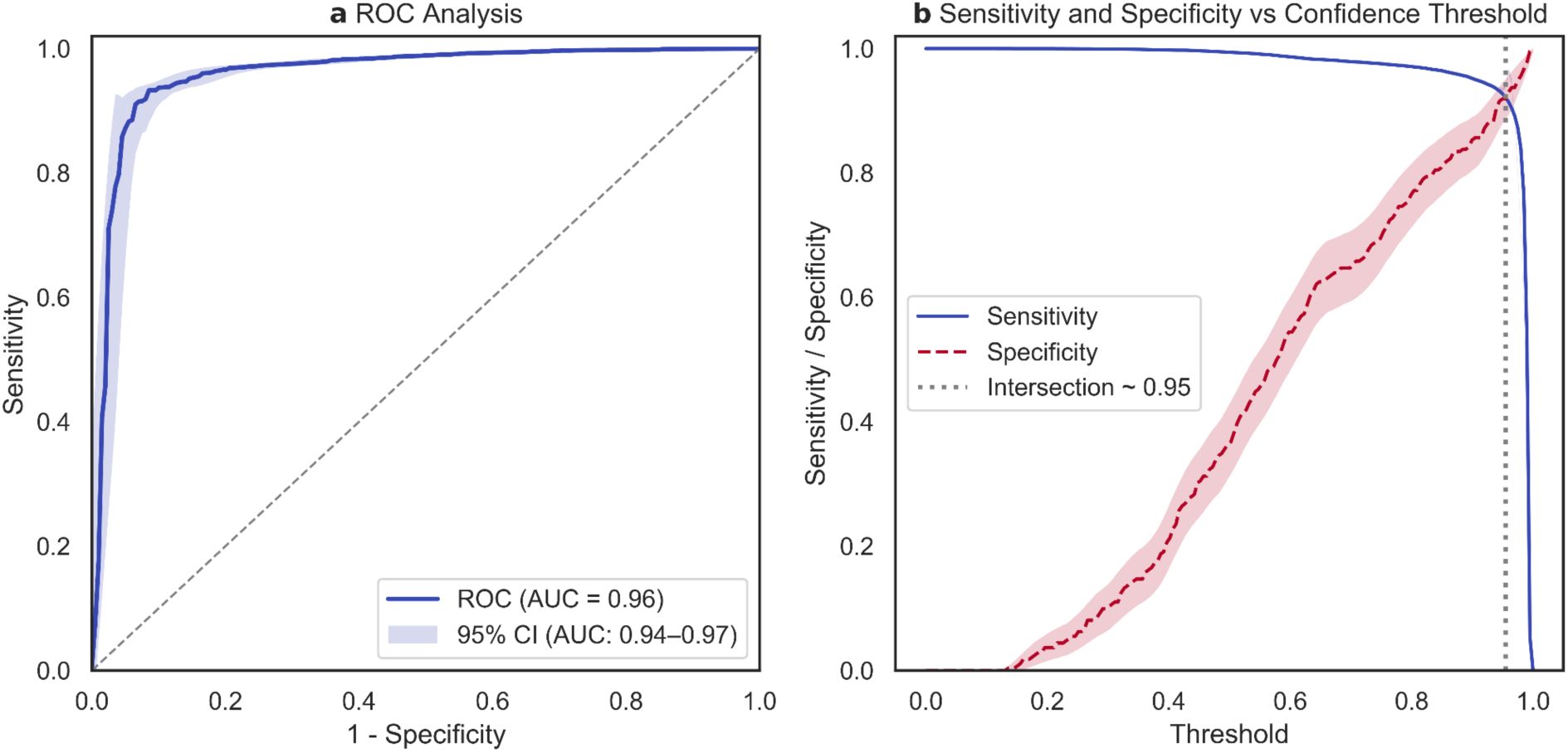
ROC analysis and confidence threshold determination on scorable samples (n=16,398, tumor and normal) **a**, ROC analysis on aggregated scorable samples. **b**, Sensitivity and specificity (y-axis) changes based on confidence threshold (x-axis), showing optimal threshold for high confidence predictions at ∼0.95. ROC – receiver operating characteristic, AUC – area under the curve, CI – confidence interval.

### Dual class B-ALL samples

For exploratory analysis, we investigated the performance on tumors having multiple hallmark molecular markers. For this, we used pediatric B-cell acute lymphoblastic leukemia (B-ALL) samples from St. Jude’s cohort^11^ with two B-ALL class-defining markers. These samples were excluded from the training cohort. We then evaluated the top three predictions by FRANK and evaluated whether they captured both molecular markers. At least one B-ALL marker was detected for all such cases, and both markers (namely, *BCR::ABL1*/*BCR::ABL1*-like rearrangements, *CRLF2* rearrangements, *ETV6::RUNX1/ETV6::RUNX1*-like rearrangements, intrachromosomal amplification of chromosome 21 (*iAMP21*), *PAX5* rearrangements, and hyperdiploid karyotype) were detected in 44/61 (72.1%) samples (Fig. 4). Interestingly, some combinations, notably *PAX5* alteration with hyperdiploidy, were challenging to detect with only hyperdiploidy being detected, but not *PAX5* alterations. These results suggest that while the majority of the class-defining abnormalities contribute to the transcriptomic changes in a complementary manner, some of the classes display dominant behavior over others.

**Fig. 4:**
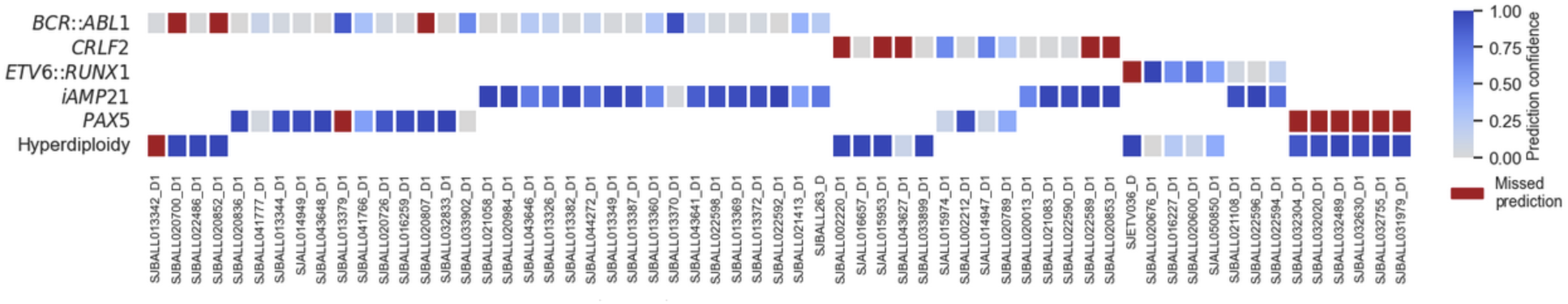
FRANK predictions on dual class B-ALL cases. X-axis lists the IDs of samples with two hallmark molecular B-ALL aberrations from the St. Jude cohort. Y-axis lists the molecular aberrations. Red squares correspond to predictions missed by FRANK. Grey-to-blue squares are correct predictions where one of the top three FRANK predictions corresponded to one of the two molecular changes. The color of grey-to-blue squares corresponds to the confidence score for that prediction. BCR::ABL1 – BCR::ABL1 and BCR::ABL1-like rearrangement, CRLF2 – CRLF2 rearrangement, ETV6::RUNX1 – ETV6::RUNX1 and ETV6::RUNX1-like rearrangement, iAMP21 – intrachromosomal amplification of chromosome 21, PAX5 – PAX5 rearrangement.

### Robustness analysis

Next, we evaluated the robustness of FRANK. First, we simulated gene dropout at different fractions and evaluated FRANK on the combined RCCUH dataset (*n=*189, tumor and normal samples). As expected, the performance decreased with increasing fraction of dropped genes. However, only a 10% reduction in accuracy was only observed when 1,921/5,260 (36.5%) or more genes used for input were missing, showing model robustness for missing data (Fig. 5a). Increasing entropy (noise) of gene expression yielded a similar result (Fig. 5b). Notably, the effect of rounding input data or reducing the amount of information using bit quantization was well tolerated up to rounding to whole numbers (Fig. 5c) and 4-bit quantization (Fig. 5d), showing robustness to different data processing. Finally, performance remained almost identical when tested on different GENCODE versions (v31 and v29) (Extended Data Fig. 1h). In summary, this highlights that FRANK is robust for different data variation sources, including sparsity, noisiness, compression, and GENCODE versions.

**Fig. 5:**
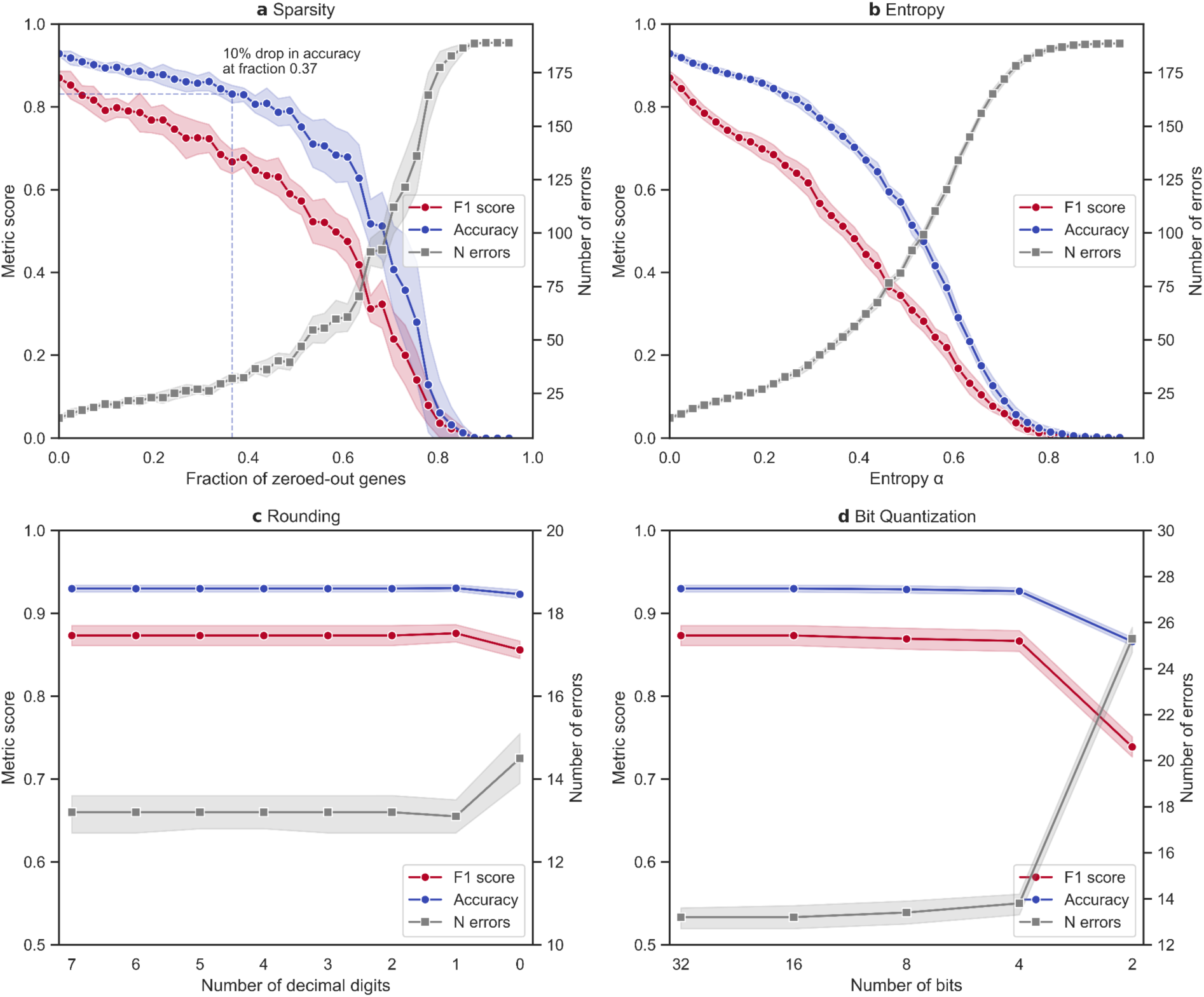
Robustness analysis on combined RCCUH dataset (n=189, tumor and normal samples) **a**, Effect of gene dropout (sparsity) on FRANK’s performance. X-axis represents the fraction of genes for which the values are set to 0 prior to model input. **b**, Effect of increasing entropy (noise) on FRANK’s performance. X-axis represents the fraction of genes for which noise is introduced prior to model input. **c**, Effect of rounding input on FRANK’s performance. X-axis represents the number of decimal digits in the input values. **d**, Effect of quantizing input on FRANK’s performance. X-axis represents the number of bits that is used to quantize input values. In **a**, **b**, **c**, and **d**, y-axis on the left shows accuracy and F1 score in the range 0–1, y-axis on the right shows the number of errors.

### Effect of tumor purity

To further investigate the robustness of FRANK in various clinical settings, we analyzed the effect of tumor purity on its performance. Detailed purity data were available for 158 (88.8%) of 178 RCCUH tumor samples (Supplementary table 17). For liquid tumors, purity was determined using flow cytometry, whereas for solid tumors it was estimated using PureCN^24^ and variant allele frequencies of driver mutations. The performance metrics of FRANK appeared to be reasonably comparable between different tumor purities, with accuracy ranging from 83.3% for tumors with 37.5–50.0% purity to 100% for tumors with 50.0–75.0% purity (Extended Data Fig. 3). Given that different tumor types have different purities, we have performed independent analysis using *in-silico* dilution using combinations of tumor and corresponding normal tissues (Methods). This revealed that performance metrics were optimal for the simulated samples with tumor purity ranging from 37.5 to 100% (Extended Data Fig. 4).

### Long-read RNA-seq data

To assess FRANK’s performance on out-of-distribution data, we tested it on three datasets sequenced using Oxford Nanopore Technologies (ONT). The first ONT dataset included 23 (15/23 (65.2%) patients ≤18 years old) osteosarcoma (fine-level class) samples and 13 normal tissue samples^25^. Of these, 18 (78.3%) were correctly identified as osteosarcoma and all 13 (100%) were correctly identified as non-malignant samples by FRANK. Incorrect predictions included synovial sarcoma and Ewing sarcoma, as well as three samples were predicted as non-malignant tissue (one muscle and two skin) (Table 4), suggesting low tumor purity in these samples, although purity was not assessed in the original study. The second ONT dataset included 12 adult cases of clear cell renal cell carcinoma^26^, all of which (100%) were predicted correctly (Table 4). Finally, we tested FRANK on 87 GTEx samples (all adult subjects) obtained using long-read ONT sequencing (GTEx LR)^27^, of which only a single sample was classified incorrectly (accuracy 98.9%, brain tissue predicted as heart). There was no overlap between GTEx patients used for training and GTEx LR. Overall, these results show that FRANK is robust to different data types and also performs well also on long-read samples despite being trained on short-read data only.

**Table 4:**
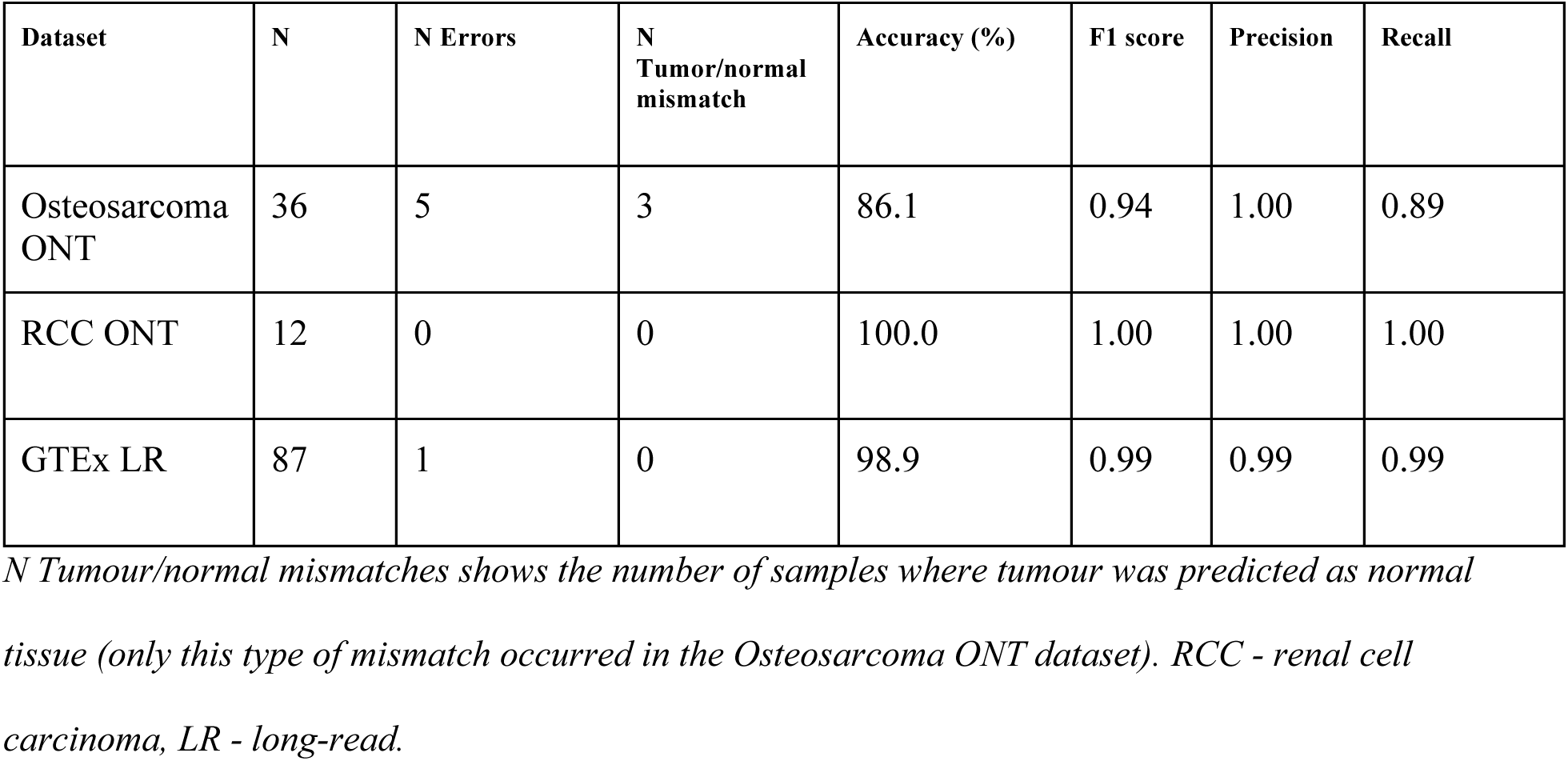
Performance of FRANK on ONT data.

### Clinical utility

To test the clinical utility of FRANK, we have classified five samples from the RCCUH cohort that did not have a definitive subclass (corresponding to fine-level) classification after comprehensive clinical testing (classified as not-otherwise-specified (“NOS”)) (Supplementary table 16). FRANK predicted a fine-level class with high confidence (>0.95) for two of these samples. The first sample, initially classified as B-ALL NOS, was classified as *PAX5*-altered B-ALL (0.985 score). Upon manual review of the sequencing data, an in-frame *PAX5* exon 5 internal tandem duplication (*PAX5*-ITD) was identified (Fig. 6a). The isolated exon 5 ITD is an extremely rare variant type in the *PAX5* gene^28^. It was initially classified as a variant of uncertain significance because it is located outside of known mutational hotspots, and the functional effect of the ITD is not known.

**Fig. 6:**
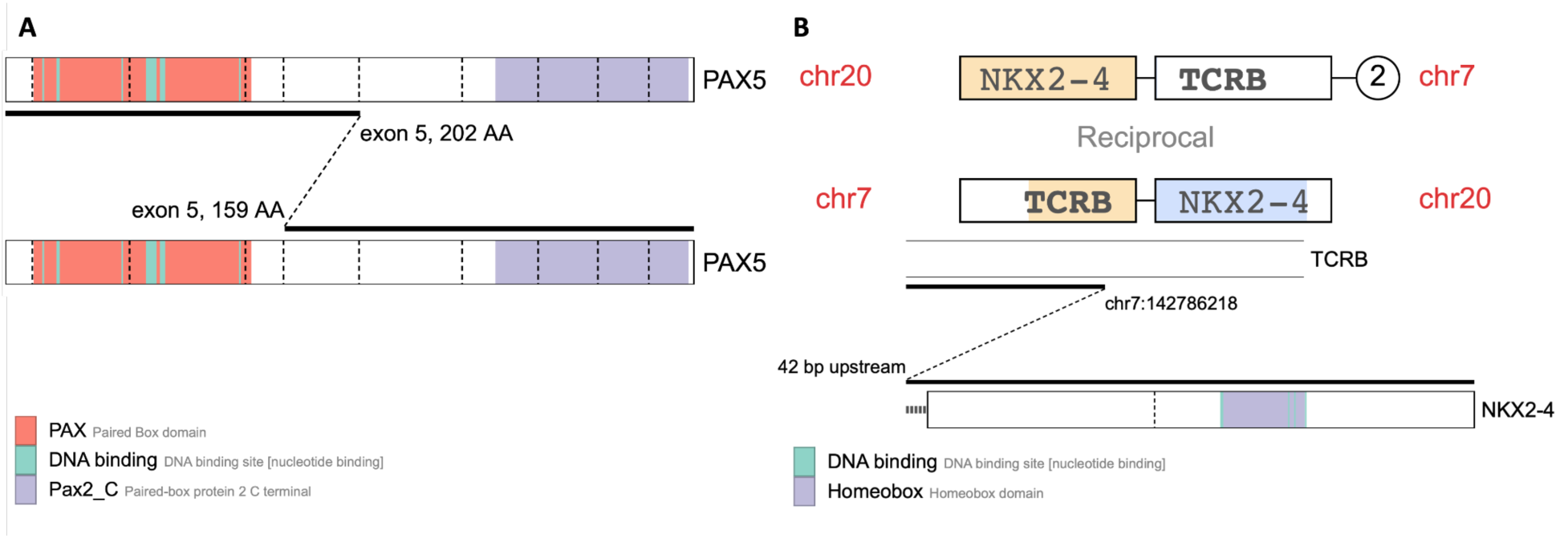
Clinically actionable variants prioritized after FRANK classification. **a**, Visualization of PAX5 exon 5 internal tandem duplication **b**, Visualization of the reciprocal t(7;20) forming TCRB::NKX2-4 fusion. Visualizations made using CICERO^36^.

The second sample initially classified as T-ALL NOS, was classified as *NKX2*-activated T-ALL (0.980 score), typically caused by translocation between T-cell receptor (TCR) genes and *NKX2-1* or *NKX2-5* genes causing overexpression of the respective *NKX2* genes via enhancer hijacking^29^. Upon reanalysis of the sequencing data, a reciprocal translocation t(7;20) forming a hybrid gene between the *TCRB* and *NKX2-4* genes was identified. The *NKX2-4* gene is currently not confirmed to be an oncogene in OncoKB^30^ or COSMIC^31^ and fusions with *NKX2-4* were not identified among 1309 T-ALL pediatric cases^12^, so the fusion was not prioritized during the initial analysis. During the reanalysis, *NKX2-4* was found to be the most overexpressed gene in the sample (Z score=341, p_adj_<0.001 when compared to other hematological samples), confirming the functional consequences of the fusion. Additionally, we were able to identify four cases with similar *NKX2-4* fusion in the literature likely confirming the role of NKX2-4 fusions in the development of the NKX2-activated T-ALL^32–35^.

For the three other samples, no additional variants were identified upon reanalysis and all predictions were with low confidence (0.67, 0.38 and 0.28, respectively), suggesting that these cases represent rare (potentially not yet described) classes. In summary, FRANK facilitated the identification and interpretation of variants in samples that had previously been devoid of a definitive subclass.

### Unknown classes

We further evaluated FRANK’s performance on unknown classes by testing it on 15 RCCUH samples with known rare tumor subclasses which were not present in the training set (Supplementary table 16). Only for 1(6.7%) of the 15 samples FRANK had predictions with high confidence (>0.95) and classified the sample with final diagnosis of Spindle Cell Rhabdomyosarcoma as Embryonal Rhabdomyosarcoma. This shows that FRANK does not assign high confidence to unknown classes, and that prediction scores can be utilized to guide further diagnostic investigations.

## Discussion

In this work, we introduced a highly accurate and robust neural network model FRANK for classifying pediatric tumors based on transcriptomic (RNA-seq) data and performed extensive benchmarking on various datasets. We demonstrated that FRANK offers significant improvement when compared with previously described methods/tools in terms of accuracy and available classes, and furthermore provides clinical utility for pediatric tumor diagnostics.

We observed that FRANK’s high accuracy was achieved via several important steps. Firstly, we performed comprehensive sample curation and alignment across training sets according to established and emerging molecular disease classification. This not only improved the performance of FRANK, but also enriched data for rare disease entities. While OTTER has a higher number of total classes, its class definition depends on clustering results and is not necessarily supported by existing classifications, such as WHO or ICC.^9^ Secondly, we introduced a set of methods to improve the training of neural networks on RNA-seq data. Specifically, we implemented the use of various data augmentation techniques for training and inference, and introduced a hierarchical penalty term that encouraged tumor class hierarchy during training. Correct training sample set grouping, data augmentation and the hierarchical penalty term all contributed to crucial improvement of the model during its optimization steps. Compared to other previously described pan-cancer classifiers^9,10^, an additional important advantages in our work is the size and diversity of the validation cohorts we employed to demonstrate the generalizability of the model.

Quantifying FRANK’s performance accurately was challenging as molecular data and tumor purity were not available for most of the datasets. Therefore, some of the misclassified results could be attributable to low purity samples or incorrectly assigned (emerging) molecular class. For example, Beat AML cohort^21^ showed lowest accuracy (85.1%) across the tested validation cohorts, which we believe may be due to the fact that it included adult samples and also had a high proportion of myelodysplastic syndrome samples, which typically have tumor purity <10%^37^. However, when benchmarking on the well-characterized external datasets (RCCUH and Princess Maxima hospital’s dataset^10^), FRANK outperformed other pediatric cancer classifiers as M&M and OTTER, achieving correct classifications for the vast majority of samples.

To demonstrate the generalizability of FRANK in various clinical settings, we tested it on multiple external datasets spanning different enrichment methods and GENCODE versions. In addition, we showed that FRANK performed well on long-read sequencing data and was robust on noisy data. Finally, we publicly release the model weights as open source and also publish a DNAnexus app for public use. In fact, based on the results presented here, FRANK has already been implemented into the transcriptome bioinformatics pipeline for clinical diagnostics at RCCUH.

We envision several different contexts in which our model will prove useful. First, FRANK can be used to guide analysis of genetic data. As shown with the two clinical cases devoid of a definitive subclass from the RCCUH cohort, our model enabled the correct classification of specific aberrations which otherwise were misinterpreted or missed in standard analysis. With whole-genome sequencing becoming increasingly available^3,38^, prioritizing variants will become even more complicated^12,39,40^. FRANK represents a tool to guide such prioritization and variant interpretation. Second, we intentionally chose a lightweight model architecture with a limited number of input genes to facilitate its use in resource-constrained and time-sensitive settings. The model can be run on most modern laptops without the need of a graphics processing unit. This, coupled with the ability to tolerate gene dropout, allows it to be run in real time, while data are produced, e.g., using ONT sequencing. Finally, FRANK can detect molecular classes even without a known driver mutation, thus reducing the number of entities with “NOS” and providing personalized treatment options for more patients. Importantly, the identification of specific transcriptomic signatures may have direct therapeutic implications. In particular, a novel drug class – menin inhibitors – has demonstrated potent antileukemic activity in leukemias characterized by *HOXA/MEIS1* overexpression, leading to the approval of the first-in-class menin inhibitor revumenib for relapsed or refractory acute leukemias harboring *KMT2A* rearrangements or *NPM1* mutations^41^. Beyond these currently approved genomic indications, HOX activation class may also predict responsiveness to menin inhibition, a concept of particular relevance for patients who lack established targetable alterations^41–43^. We anticipate that FRANK will be a vital tool in the recruitment of such potential cases, once additional validations have been conducted.

This work also has several limitations. First, even though the spectrum of tumors captured is wide, not all categories are covered. For example, a common subtype of AML, especially in adults, AML with myelodysplasia-related changes^44^, was excluded from training due to multiple genomic changes leading to this disease subtype. Second, the diagnostic performance of certain disease categories could not be determined for the final trained model due to cases with these diseases not being available in the testing cohorts, for example, T-ALL with *MEF2* rearrangement. Third, the majority of samples in the testing datasets were fresh/stabilized, raising the possibility that the model’s performance on FFPE samples could be lower^45^. Finally, direct comparison of the model with M&M and OTTER was complicated by the fact that these classifiers were trained on data generated by a different GENCODE version to the current work, as well as weights not being publicly available for the OTTER model.

In summary, we provide a robust open-weight model for pan-cancer pediatric tumor classification that is validated on multiple external cohorts and achieves a higher performance compared with available state-of-the-art classifiers.

## Methods

### Training data curation

To ensure consistency in training and classification, sample type classification was aligned across the studies used. Each sample type was then assigned to a three-level hierarchy (broad, medium and fine). Broad-level classification included brain samples, solid tissue samples and hematological samples.

Medium-level classification (*n=*48) included respective histological sample types (e.g., AML), while fine-level classification (*n=*181) was based on the accepted or emerging (molecular) tumor classification as defined per WHO v5^46^ and/or International Consensus Classification (ICC)^47^ or other tumor classifications^4,48,49^.

For acute leukemias, we specifically excluded morphologically defined classes (e.g., acute myelomonocytic leukemia), as they can harbor a variety of molecular aberrations and usually do not have specific transcriptomic signatures^47^, and focused on enriched classification based on molecular changes, adding emerging disease subtypes such as AML with *PICALM::MLLT10* fusion^44^. Where available, fine-level classification was manually curated based on the class-defining molecular markers. The exact class hierarchy is listed in Supplementary Table 3. Only samples with fine-level classification were utilized for the classifier training and testing.

### Quality control in training sample selection

Filtering of training samples was performed in two main steps (Extended Data Fig. 5). First, raw counts were used to calculate the percentage of expression from the top five expressed genes for each sample (except for AALL0434 where no raw count data were available). Hematologic, solid and brain tumors (for GTEx, we additionally split pancreas and pituitary due to batch effect) were then analyzed separately for each training dataset and samples with top five gene expression percentage above the 99th percentile were filtered out to reduce biased TPM across samples^50^.

Next, for each dataset and tumor type, the groups were further subdivided, when available, based on the enrichment method (poly-A or total RNA). For each subgroup principal component analysis (PCA) was then performed on the TPM-normalized counts across all available genes. Finally, samples were projected onto a 2D PCA plot, the center was defined as the mean of principal components 1 and 2. Samples projected >3 standard deviations away from the center and having top five gene expression above 20% were filtered out (Supplementary Fig. 1,2,3,4). Samples excluded in QC are listed in Supplementary table 10.

### Training data and hold-out splits

The exact strategies used to select samples for training and validation across the four training datasets (St. Jude^11^, Princess Maxima^10^, AALL0434^12^, and GTEx^13^) are shown in the Extended Data Figure 6. First, QC was applied to all datasets to reject low-quality samples. Then each dataset had an individual strategy. No St. Jude samples were used for performance metrics testing, so all samples remaining after filtering strategies were used for training. Data for St. Jude samples were downloaded from the St. Jude Cloud up until October 2025.

For testing on the Princess Maxima dataset, we first selected the same samples used for testing in the original publication (*n=*472)^10^. Of these, 446 (94.5%) could be assigned to a fine-level class in FRANK, while the rest (*n=*26, 5.5%) had no specific fine-level class (e.g., “B-ALL, NOS”) or did not have a matching class (*n=*26; 5.5%).

For the AALL0434 dataset, “NOS” categories were first removed. Then, for each remaining category we randomly selected 30% of the samples to be used for training and 70% for the hold-out testing. For example, there were 32 T-ALL *PICALM::MLLT10* samples in AALL0434, of which nine were assigned to the training set and 23 to the test set.

Regarding the GTEx dataset, samples were split by donor IDs (given that one donor had multiple tissues): 30% of donor IDs for training and 70% for hold-out testing. Then, the samples were moved to training or testing based on the donor ID, thus avoiding samples from the same donor being in the training and in the testing sets.

All training samples were generated using short-read sequencing data, transcriptomes were obtained using a mixture of fresh/frozen samples and FFPE, as well as poly-A and total RNA enrichment. We intentionally chose not to use TCGA^51^ cohorts for training as they contain tumors that do not generally represent the pediatric population and typically lack fine-level classification information.

### Transcriptomic data curation

We reviewed the fine-level tumor subtypes for the training and testing datasets using provided sample classification and molecular drivers (where available) to ensure data consistency. Data curation strategy for datasets shared between training and testing datasets (Princess Maxima, GTEx, and AALL0434) is described in section “Training data and hold-out splits”. For TARGET AML, we reviewed the molecular findings from the original publication^15^ (Supplementary tables S5 and S6 in the publication). TARGET Other classes corresponded to the fine-level classification. Beat AML samples were harmonized by mapping classes provided in clinical data (accessed through University of California Santa Cruz Xena Browser^52^. Saint-Louis cohort ground truth classes were extracted from Gu et al.^53^ (Supplementary table S5 in the publication).

In all datasets, cases that did not have a fine-level subclass or definitive molecular finding were labeled as “NOS” and were not used in the study.

### Riga Children’s Clinical University Hospital Cohort

All pediatric tumor samples from the Riga Children’s Clinical University Hospital, Latvia (RCCUH) cohort had undergone deep (>250×) exome sequencing for single nucleotide variant/indel and copy number alteration (CNA) detection, as well as deep transcriptome sequencing for fusion and overexpression analysis, as a part of the hospital’s standard molecular tumor work-up. Where required, additional testing was performed according to the respective treatment protocols. For brain tumor samples without a clear histological or molecular diagnosis, DNA methylation-based testing using the Heidelberg CNS Tumor Methylation Classifier^4^ was performed via outsourcing.

Tumor samples were stabilized using PAXgene tubes (for liquid tumors) or Nucleoprotect reagent (for solid tumors) prior to RNA extraction. RNA samples with high fragmentation (DV200 < 30%) were not processed. Transcriptome sequencing was performed at CeGaT, Tübingen, Germany, using KAPA RNA HyperPrep with RiboErase (HMR) with KAPA Globin Depletion Hybridization Oligos (Roche, Basel, Switzerland) for library preparation. Libraries were sequenced as 100 PE on an Illumina NovaSeq X Plus platform, generating at least 100M clusters per sample.

The deep transcriptome data processing was performed on the secure DNAnexus platform. Reads were mapped using STAR aligner v2.7.9a^54^ and analysed using CICERO v1.9.6^36^, STAR-fusion v1.11.0^55^, and arriba v2.3.0^56^ tools, with fusion call merging using EnFusion v1.0.0^57^. Reads were counted using HTSeq^58^ and normalized as TPM. Expression outlier detection was performed by comparing a sample’s TPMs to the other samples from the same high-level classification using t-test and OutSingle v1.0.0^59^.

### Model training

#### Input feature set selection

Merged TPMs for the four training datasets contained 45,698 transcript types. From these, we first selected the following transcripts based on HUGO Gene Nomenclature Committee (HGNC)^60^ symbol types: gene with protein product, RNA, long non-coding, immunoglobulin gene, and T cell receptor gene. This yielded 13,349 transcript types. Next, using the training dataset with these transcripts, we trained an RF classifier using a three-fold stratified split (corresponding to the number of samples for the most underrepresented classes). We then selected the top 7,000 genes ranked on average importance across the folds (Supplementary table 4). Finally, TPM values for the 7,000 genes were *log_2-_*normalized and filtered using a minimum variance threshold of 1.0 resulting in a total of 5260 genes used as the input for the model training (Supplementary table 5).

#### Neural network architecture

The final trained model was a three hidden layer fully connected neural network with batch normalization, leaky rectified linear unit (ReLU) activation and dropout in each layer. The model produces separate softmax-normalized probability distributions for broad, medium, and fine classes. Predictions for the broad, medium and fine levels are made from the first, second and third hidden layers, respectively.

#### Data augmentation

To achieve equal performance across different datasets and tumor types, we performed extensive data augmentation. Rare classes (n≤5) were augmented by averaging gene expression from possible sample combinations, yielding in new synthetic examples. This simple strategy was chosen because techniques such as Synthetic Minority Oversampling Technique (SMOTE) require a higher number of examples^61^. During training, we also applied augmentations including random feature dropout and noise, global and feature-level noise, global and feature-level scaling, and CutMix^14^ to account for the different number of features present in different GENCODE versions and provide robustness of the classifier for data missingness.

#### Hierarchical regularization

High-level predictions can inform more accurate fine-level predictions. To account for tumor hierarchy in the training and enforce consistency between predictions at different levels of the label hierarchy (broad, medium, fine), we used a hierarchical regularization during training. We first defined fixed binary mapping matrices that encode valid parent–child relationships across hierarchy levels. Then, for each sample, we computed the probability mass that was outside the allowed regions. The final hierarchical regularization term was a weighted sum for broad-medium, medium-fine and broad-fine penalties.

#### Neural network training and inference

We utilized separate cross-entropy losses for the three hierarchy levels, with equal weights for all. The final training loss was the sum of the hierarchical regularization term and the three cross-entropy losses. We used the Adaptive Moment Estimation (Adam)^62^ optimizer with a constant learning rate and a batch size of 32. Training of the neural network was run for 350 epochs with patience set to 150 (to avoid early stopping) on all training data, selecting the epoch with the best F1 score for fine-level class as the final trained model. Since the training used CutMix augmentations, no overfitting was observed even after training for a higher number of epochs. To test this hypothesis, we trained the model with CutMix turned off and observed a training curve consistent with overfitting (Supplementary Fig. 5). Reproducibility during training was ensured by setting random state to 42.

Inference was performed using 20 iterations of test-time augmentations (excluding CutMix), averaging the logits and selecting the class with the highest mean prediction score. If input for one of the genes was missing, it was set to 0.

#### Other machine learning model training

RF, LR, and SVM were trained using the default settings. XGB was trained with default settings, except that the evaluation metric was set to “logloss”. Reproducibility was ensured by random state 42. The training dataset consisted of the same samples used to train the neural network. The same pipeline of log2 normalization, variance filter, and standard scaling was applied to the inputs when using these models.

### Benchmarking

Benchmarking was performed against two published pan-cancer RNA-seq classifiers: M&M^10^ and OTTER^9^. Different classifiers have different sets of classes that they predict and sometimes do not have direct mapping between them. In some cases, predictions could be considered correct at a hierarchical level, for example, if the ground truth was “AML with *NPM1* mutation” but the predicted class was “AML”.

#### Comparison to M&M

Comparison to M&M was performed on scorable datasets that used short-read sequencing: RCCUH, RCCUH-29, Princess Maxima, RSU, TARGET AML, AALL0434, TARGET Other, Saint-Louis T-ALL cohort, and Beat AML. Since M&M has a limited number of non-malignant classes, comparisons with GTEx were not performed. We first mapped the unique fine-level categories in these datasets to available M&M’s fine-level prediction classes (Supplementary table 6). We then selected only the samples that could be mapped to M&M’s fine level class (Supplementary table 7). Since M&M only has a class for non-malignant bone marrow but not for non-malignant whole blood, we merged non-malignant bone marrow and whole blood into a single class for this analysis. M&M’s predictions are provided in Supplementary table 8 (column “include_in_comparison” shows samples that were mapped to FRANK classes). Since both M&M and FRANK output top three predictions, for accuracy comparison with M&M we additionally report the top three accuracy.

M&M inference was run using R version 4.5.0 following developers instructions.

#### Comparison to OTTER

OTTER does not provide model weights and could not be run locally. Due to limited access to OTTER inference, it was compared on RCCUH samples only. The samples were run with OTTER (<u>otter.ccm.sickkids.ca/app/v1</u>) using the Model Name “otter”, choosing the deepest hierarchy level prediction. For comparison to FRANK and M&M, we first evaluated whether RCCUH fine-level categories could be mapped to deepest hierarchy level OTTER classes (Supplementary table 9, column “otter_evaluate”). For samples that had a match to OTTER classes, we mapped them to FRANK fine-level class (Supplementary table 9, column “mapped_prediction”). Only 0.63% of genes were shown as missing by OTTER during inference for RCCUH samples.

### Packages used

Model training and data analysis were performed using Python 3.12.9, virtual environment implemented with uv 0.8.11. Neural network training was performed using PyTorch 2.7.1, matrix manipulation with NumPy 1.26.4, tabular data manipulation using Pandas 2.2.3. Visualizations were generated using Matplotlib 3.10.3 and Seaborn 0.13.2. RF, LR, SVM, and principal component analysis were implemented using scikit-learn 1.6.1. XGB was implemented using xgboost 3.0.0.

### Metric calculation

F1, precision and recall for multiclass classification metrics were calculated using the *macro* averaging strategy with scikit-learn 1.6.1. Additionally, *labels* parameter was used with unique ground truth values passed. 95%CI for AUC was calculated using 2000 iterations of bootstrapping.

### Robustness analysis

We used the RCCUH dataset as a pan-cancer dataset for sparsity analysis to evaluate FRANK’s robustness. 20 test-time augmentations were used for the inference of each sample. For simulated gene dropout, a fraction of the 5260 input genes was removed by setting their values to 0. We generated 10 iterations of randomly dropped genes at each fraction using different random seeds. Incremental entropy increase was simulated by adding noise with increasing noise fraction (0 – original data, 1 – pure noise).

For each fraction, we generated 10 noisy data sets using different random seeds. The effect of decimal digits was evaluated by decreasing the number of decimal digits from 7 to 0 (whole numbers) on the log-normalized input values. At each decimal digit number, we ran 10 iterations using different random seeds for test-time augmentations. The bit-quantization effect was tested by quantizing log-normalized input with bit sizes of 32, 16, 8, 4, and 2. 10 iterations using different random seeds for test-time augmentations were used for each bit size.

### *In-silico* dilution

To evaluate FRANK’s performance on different tumor purities, we performed simulated dilution according to a previously described approach (MethaDory)^63^. We first selected a set of corresponding normal samples for each RCCUH medium-level class in samples with tumor purity available (e.g., tumor: “B-cell Acute Lymphoblastic Leukemia”, corresponding normal: “Non-malignant Bone Marrow”, Supplementary table 18). Next, we simulated tumor dilution with an elementwise sum of tumor and normal tissue TPM vectors, followed by renormalization:

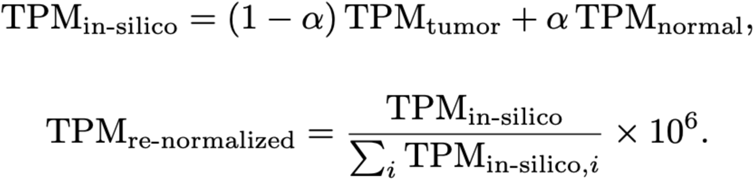

where ɑ is the fraction of normal tissue

The new tumor purity was then estimated by P*_in-silico_*=P_tumor_(1-*ɑ*). We linearly sampled 39 ɑ values in the range 0.05–0.95 and performed 10 iterations at each dilution level by randomly selecting a different corresponding normal sample in each iteration. Metrics were calculated for each iteration separately.

## Data availability

Access to St. Jude Children’s Research Hospital RNA-seq data can be requested (https://www.stjude.cloud/). GTEx v10 release data for short and long-read sequencing are openly available (https://www.gtexportal.org). RSU data are available at dataverse: https://doi.org/doi:10.48510/FK2/4EMT9P. RCCUH data are not publicly available due to institutional ethical restrictions but can be requested with bioethical committee approval. Princess Maxima Center for Pediatric Oncology data are openly available in https://www.ebi.ac.uk/biostudies/arrayexpress/studies/E-MTAB-14038. Beat AML, TARGET AML, and Target Other TPM data are accessed using University of California Santa Cruz Xena Browser (https://xenabrowser.net). AALL0434 TPM data are available at https://www.synapse.org/Synapse:syn54032669/wiki/627818 (registration required) with sample classes available in https://pmc.ncbi.nlm.nih.gov/articles/PMC11611067/#SD1 Supplementary table 3. Saint-Louis Hospital T-ALL TPM data are available under accession codes GSE110633 and GSE110636. Osteosarcoma ONT transcriptome data are available under accession code GSE218035, and Clear cell renal cell carcinoma ONT transcriptome data are available under GSE241932.

## Sample anonymization

Sample identifiers used in this study are internal anonymized codes. No correspondence between these codes and identifiable patient information is accessible outside the research group.

## Code availability

Code for FRANK (local and DNAnexus versions) will be publicly available at github upon manuscript publication: https://github.com/ksablauskas/frank-classifier

## Acknowledgements

K.S. work is in part supported by the Sanguine project that has received funding from the Horizon Europe programme (agreement ID 101097026).

We thank Anna Janberga, Santa Kursite, Žanna Kovaļova, Marika Grutupa, Zelma Visnevska-Preciniece, Gunita Medne, Irena Voitoviča, Elizabete Cebura, Anna Valaine, Iveta Račko, Dace Enkure, Elina Dimina and Nora Krike for their clinical work and valuable discussions which were fundamental to the production of this article.

## Author information

K.S. and D.R. contributed equally to this work. K.S. and D.R. contributed to the conception of the idea, data curation and analysis, writing of the original draft of the manuscript. K.S., D.R., E.B.S., L.G., I.N. and A.Z. contributed to the sample curation. K.S. performed training and inference of the machine learning models. T.K., L.B. and A.V. performed bioinformatics analysis to generate and process RCCUH and RSU data. A.J. reviewed the statistical methods used. All authors critically reviewed the manuscript and contributed to the final version.

## Corresponding author

Correspondence to Karolis Sablauskas or Dmitrijs Rots.

## Extended data figures

**Extended Data Fig. 1:**
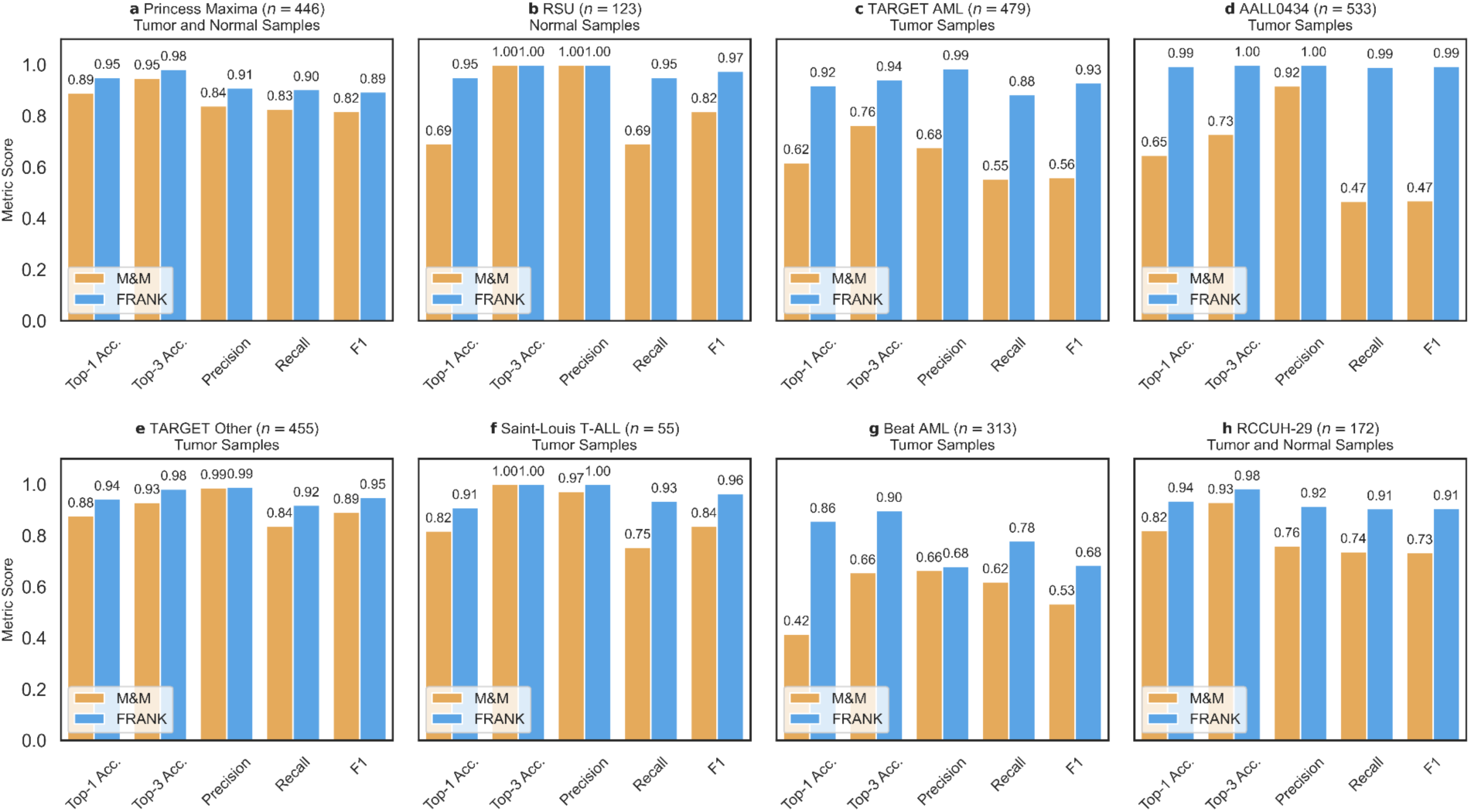
FRANK comparison to M&M on other testing datasets. **a**, Benchmarking of FRANK on Princess Maxima testing samples mapping to FRANK classes. **b**, Benchmarking of FRANK on RSU (non-malignant whole blood) samples. **c**, Benchmarking of FRANK on RSU (non-malignant whole blood) samples. M&M prediction was considered correct if it predicted non-malignant bone marrow. **d**, Benchmarking of FRANK on TARGET AML samples mapping to M&M classes. **d**, Benchmarking of FRANK on AALL0434 (T-ALL) samples mapping to M&M classes. **e**, Benchmarking of FRANK on TARGET Other samples mapping to M&M classes. **f**, Benchmarking of FRANK on Saint-Louis T-ALL cohort mapping to M&M classes. **g**, Benchmarking of FRANK on Beat AML samples mapping to M&M classes. **h**, Benchmarking of FRANK on RCCUH samples mapping to M&M classes using GENCODE v29 (RCCUH-29). All metrics are shown in range 0-1.

**Extended Data Fig. 2:**
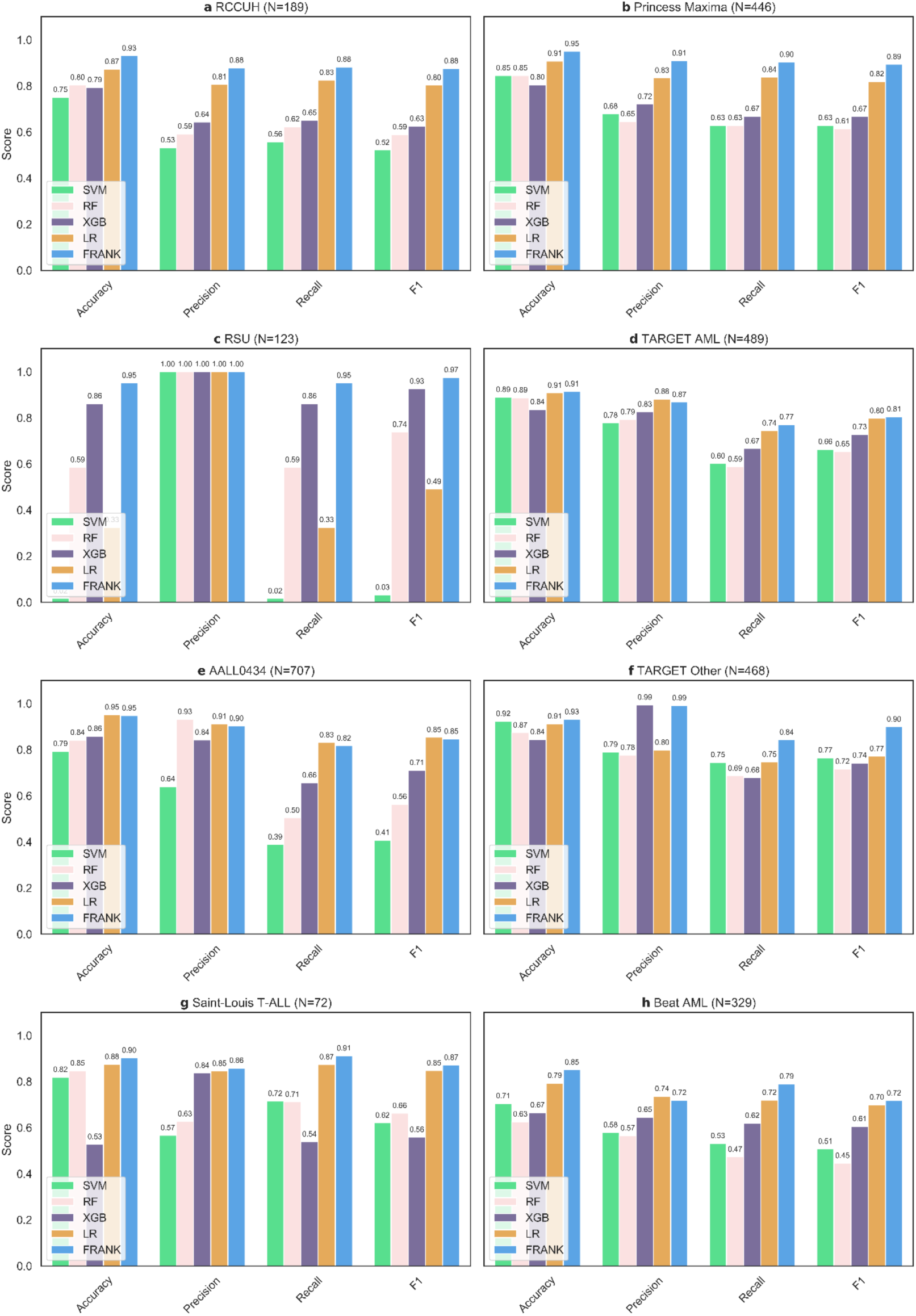
FRANK comparison other machine learning models. **a**, Performance on RCCUH dataset. **b**, Performance on Princess Maxima testing dataset. **c**, Performance on RSU (non-malignant whole blood) samples. **d**, Performance on TARGET AML samples. **e**, Performance on AALL0434 (T-ALL) samples. **f**, Performance on TARGET Other samples. **g**, Performance on Saint-Louis T-ALL cohort samples. **h**, Performance on Beat AML samples.All metrics are shown in range 0-1.

**Extended Data Fig. 3:**
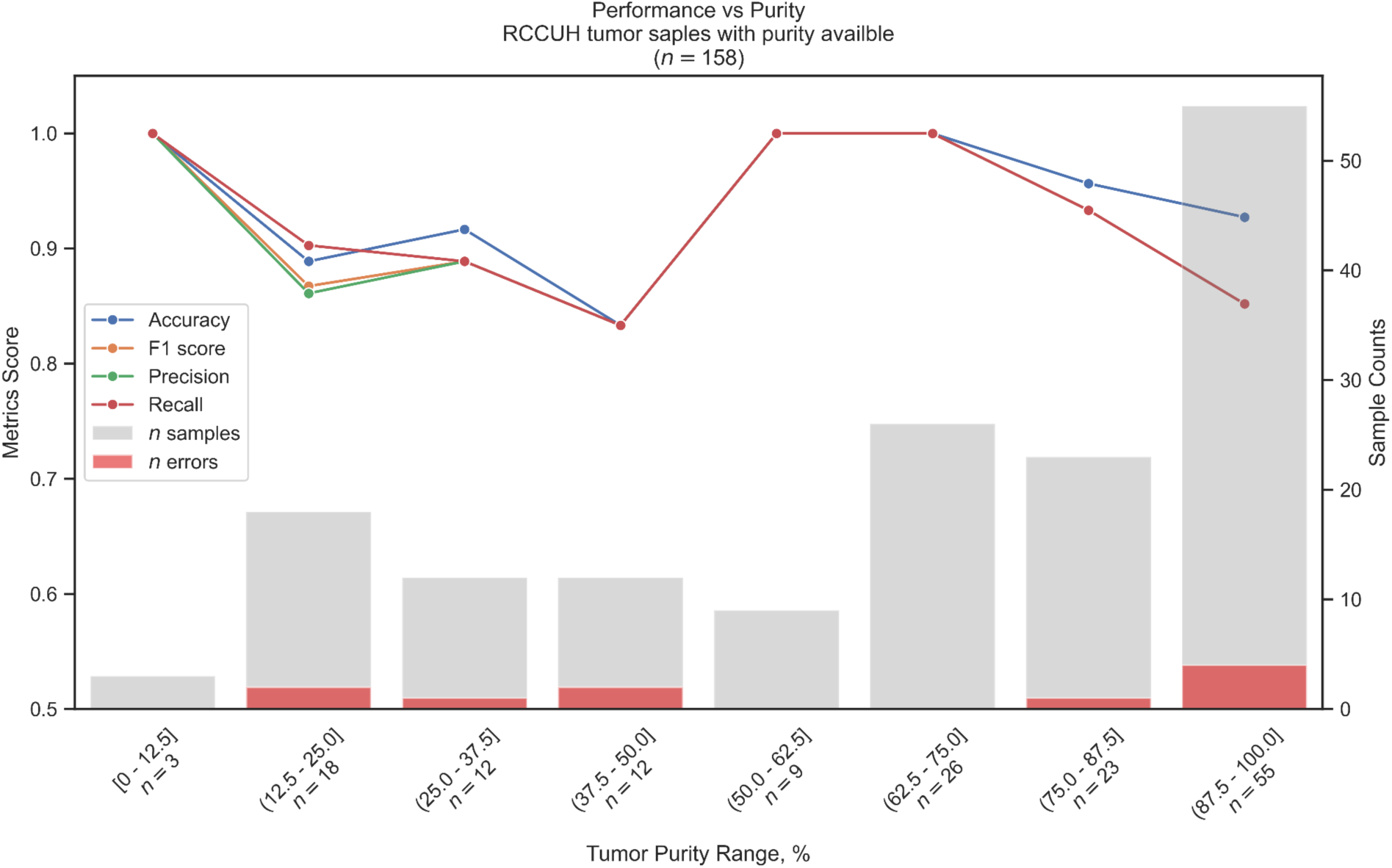
Performance based on tumor purity. The x-axis shows different estimated tumor purity ranges and the number of samples available in each bin. The y-axis on the left shows different metrics and the y-axis on the right shows total and misclassified number of samples.

**Extended Data Fig. 4:**
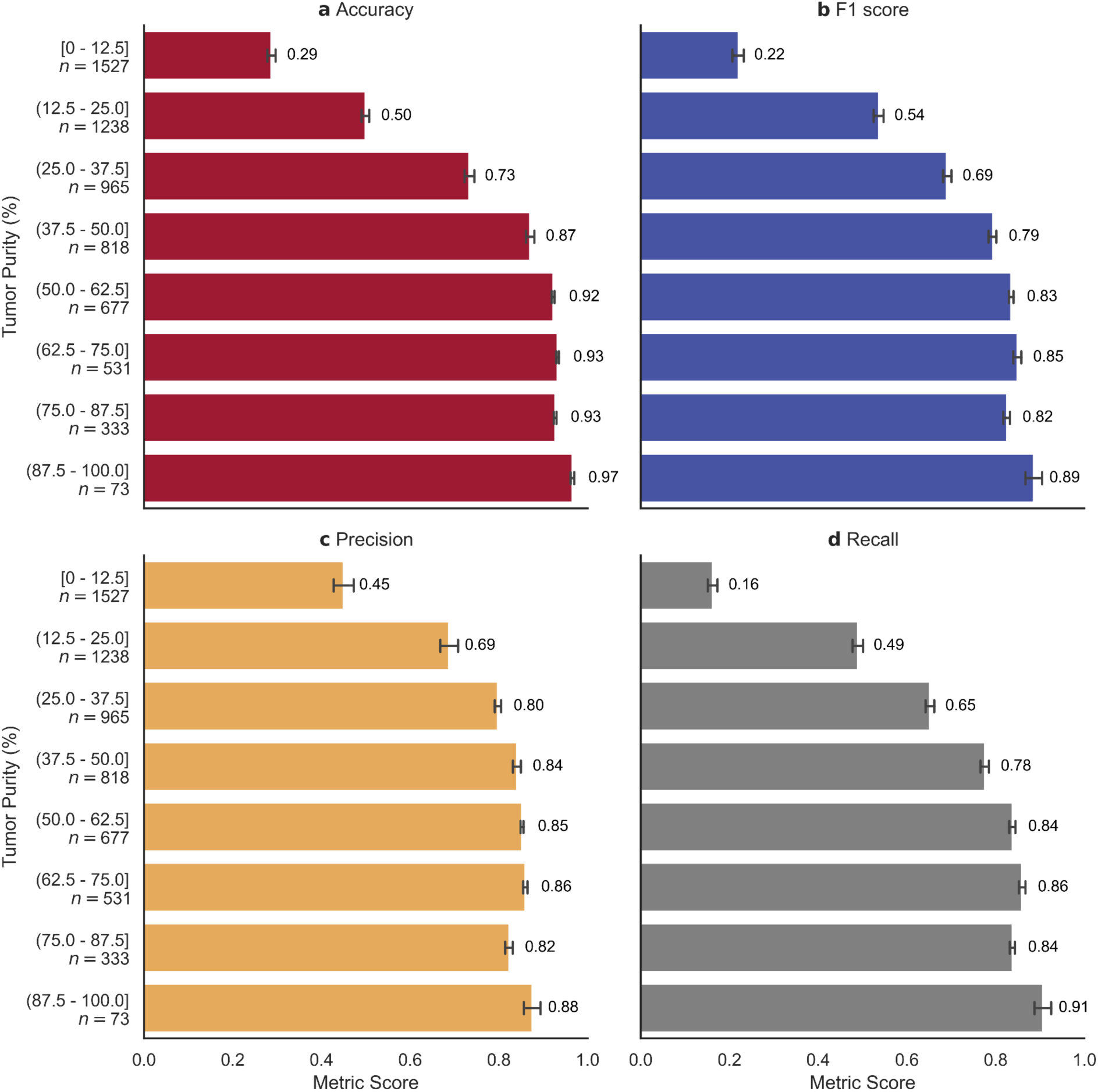
In-silico tumor dilution. The y-axis in each subplot shows tumor purity ranges and the number of samples available in each bin. The number of in-silico samples increases with decreasing tumor purity because progressively more samples with lower purity are generated (i.e. simulated purity is always lower than original purity). The x-axis in each subplot shows metrics score. The value shown is the mean performance metric across iterations, whiskers are 95%CI. **a**, Accuracy for in-silico samples. **b**, F1 score for in-silico samples. **c**, Precision for in-silico samples. **d**, Recall for in-silico samples.

**Extended Data Fig. 5:**
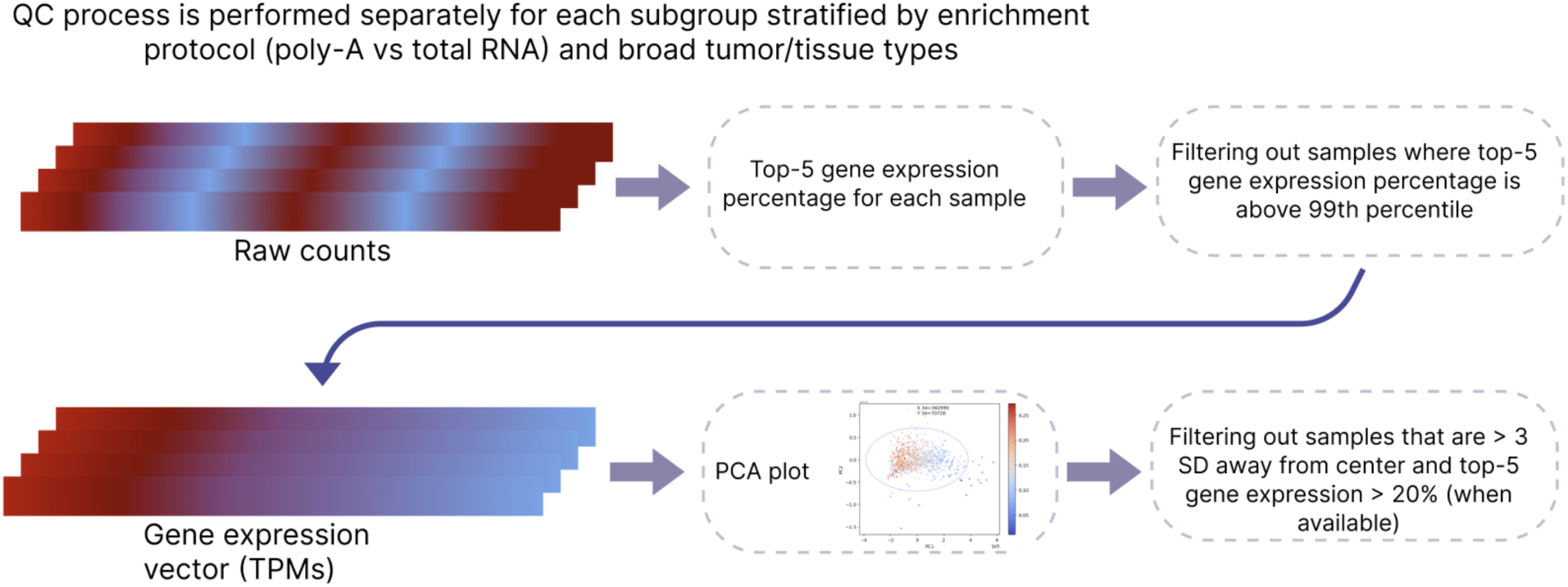
QC process Explanation of the two-step QC process. First we used raw counts (with the exception of AALL0434 where raw counts were not available) to get a top-5 gene expression percentage for each sample. Then we filtered samples where top-5 gene expression was above the 99th percentile. In the second step, we used TPMs to generate PCA plots using all genes as input. We then detect center position and draw an ellipse to detect samples that are more than 3 standard deviations away from the center. Only samples where top-5 gene expression is above 20% are filtered out.

**Extended Data Fig. 6:**
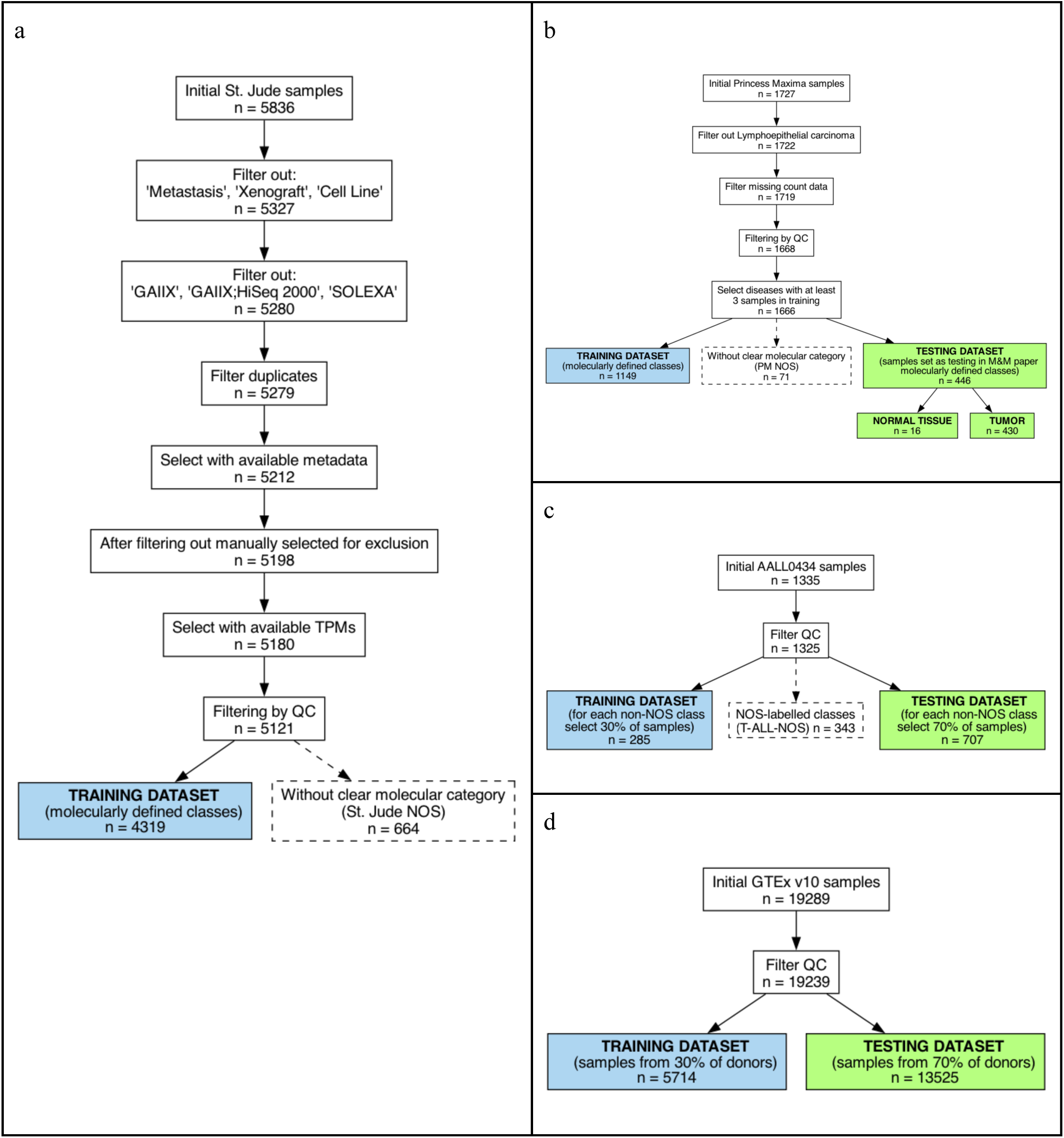
Selection of training samples and assignment to training / testing.

