## Supplementary Information for "FRANK: a pan-cancer RNA-seq classifier for childhood tumors with data-efficient learning"

### Supplementary Tables

Supplementary table 1: Training and testing numbers for each class

Supplementary table 2: All training samples with fine, medium, broad and original diagnosis

Supplementary table 3. List of hierarchy classes

Supplementary table 4: Selected genes by random forest model

Supplementary table 5: Genes used for input

Supplementary table 6: Mapping of classes to M&M

Supplementary table 7: Number of mappable samples to M&M

Supplementary table 8: M&M predictions on scorable samples

Supplementary table 9: OTTER predictions on RCCUH

Supplementary table 10: Samples filtered out by QC

Supplementary table 11: Aggregated metrics on scorable samples

Supplementary table 12: Predictions of FRANK on scorable datasets

Supplementary table 13: Predictions of FRANK on RCCUH-29

Supplementary table 14: Predictions of FRANK on double-class B-ALL

Supplementary table 15: Predictions of FRANK on ONT datasets

Supplementary table 16: RCCUH-NOS and resolved samples

Supplementary table 17: Tumor purity of RCCUH samples

Supplementary table 18: Mapping of tumor-normal for in-silico analysis

### Supplementary Figures

**Supplementary Figure 1: QC for St. Jude samples**

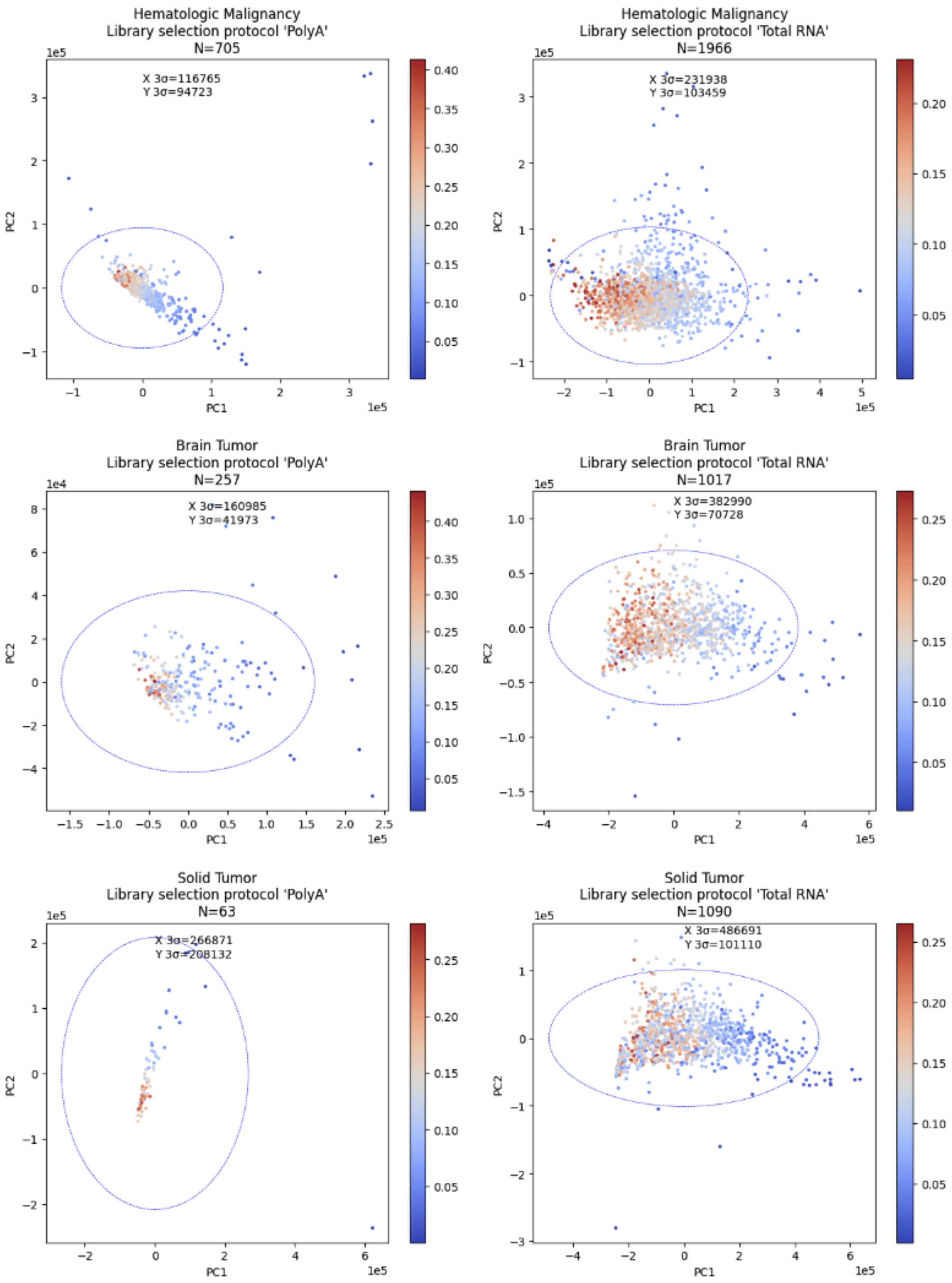

*Subplots show PCA for St. Jude samples (before QC exclusion) stratified by library selection protocol and broad sample type. Circle corresponds to 3 SD distance for PC1 and PC2 from center. Color shows top-5 gene expression ratio.*

**Supplementary Figure 1: QC for Princess Maxima samples**

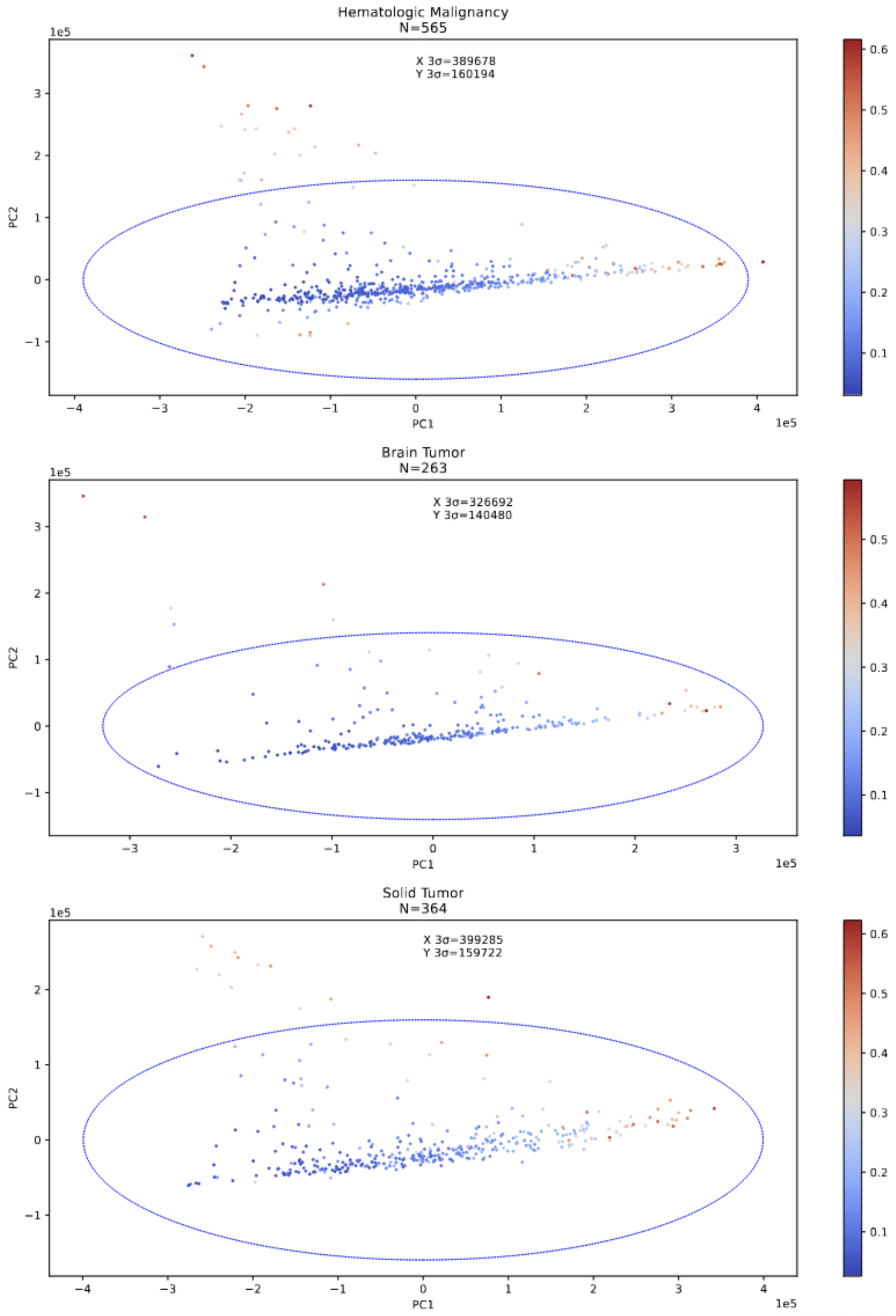

*Subplots show PCA for Princess Maxima training samples (before QC exclusion) stratified by broad sample type (all samples had the same library protocol). Circle corresponds to 3 SD distance for PC1 and PC2 from center. Color shows top-5 gene expression ratio.*

**Supplementary Figure 1: QC for AAL0434 samples**

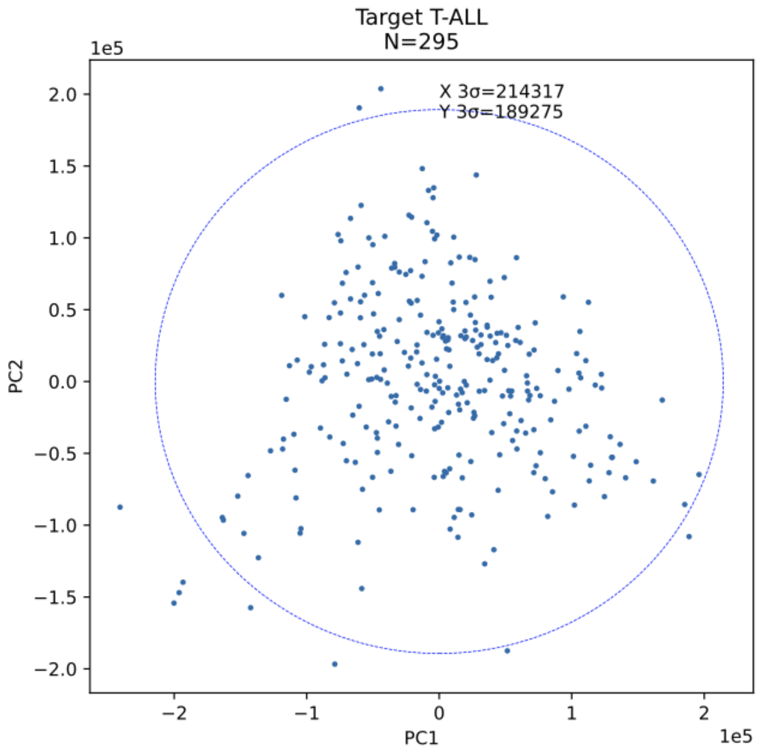


*Subplots show PCA for* AAL0434 *training samples (before QC exclusion). Circle corresponds to 3 SD distance for PC1 and PC2 from center. Color shows top-5 gene expression ratio.*

Supplementary Figure 4:

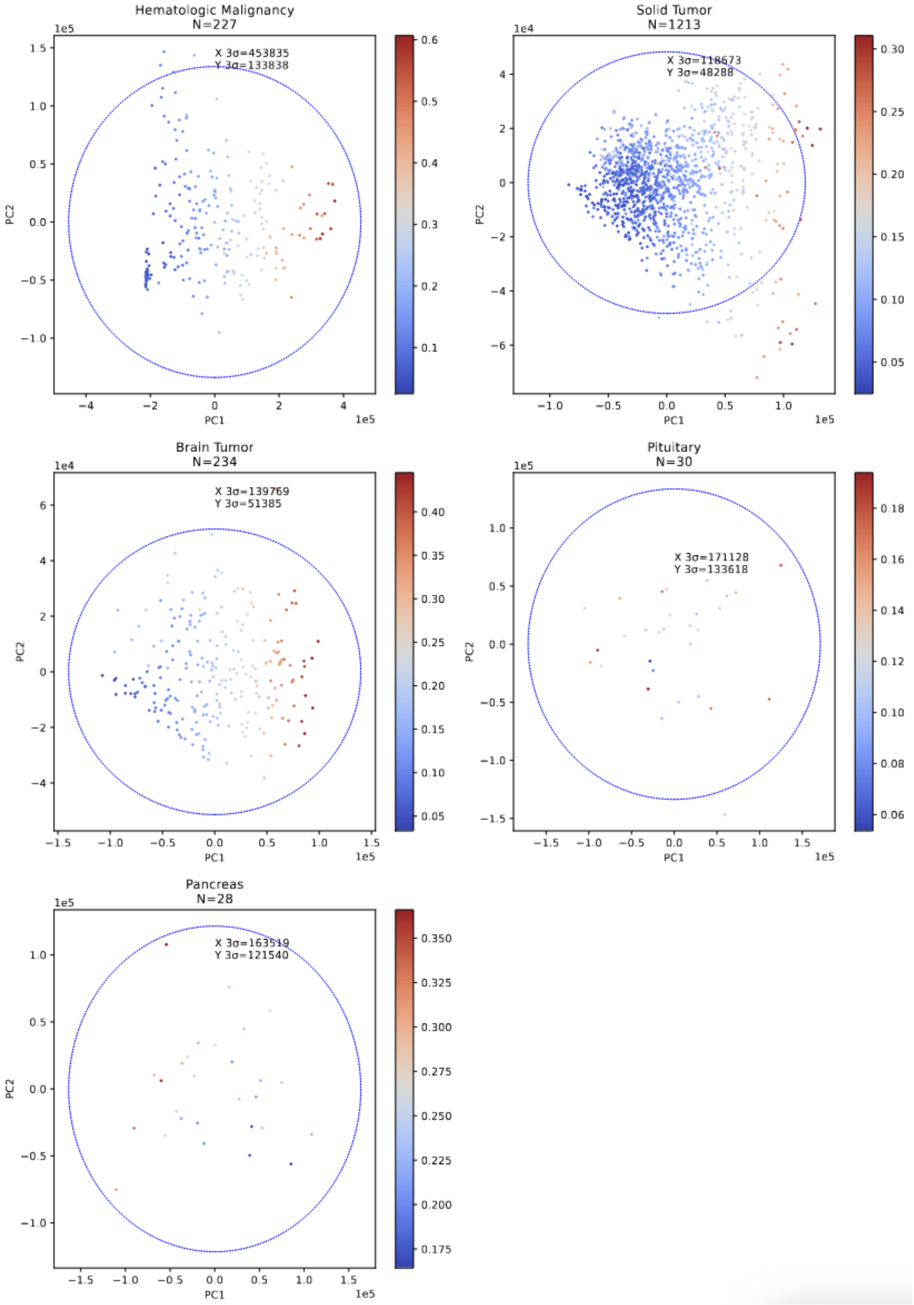


*Subplots show PCA for GTEx v10 release training samples (before QC exclusion) stratified by broad sample type (all samples had the same library protocol). Pituitary and pancreas had a batch effect and were additionally stratified. Circle corresponds to 3 SD distance for PC1 and PC2 from center. Color shows top-5 gene expression ratio.*

**Supplementary Figure 5.Training with and without CutMix**

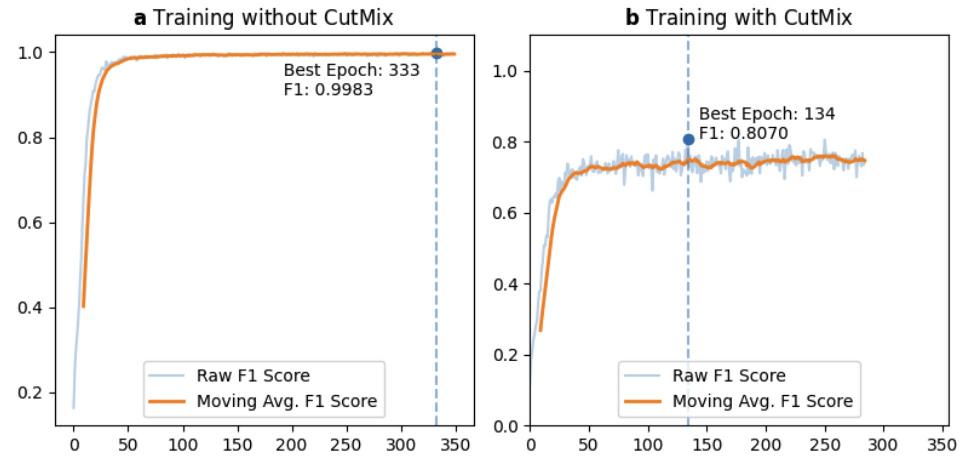


*a, Training without CutMix. The training F1 score increases and approaches 1.0 as the number of training epochs is increased which is consistent with overfitting behaviour.* ***b****, Training with CutMix. The F1 increases without apparent overfitting, despite the increasing number of training epochs. The epoch with the highest F1 score is shown.*
